# The age and sex dynamics of heterosexual HIV transmission in Zambia: an HPTN 071 (PopART) phylogenetic and modelling study

**DOI:** 10.1101/2025.05.02.25326873

**Authors:** M. Hall, W. Probert, L Abeler-Dörner, C. Wymant, F. Di Lauro, X. Xi, R. Sauter, T. Golubchik, D. Bonsall, M. Pickles, A. Cori, J. Bwalya, S. Floyd, N. Mandla, K. Shanaube, B. Yang, P. Bock, D. Donnell, M.K. Grabowski, D. Pillay, O. Ratmann, S. Fidler, H. Ayles, R. Hayes, C. Fraser, the PANGEA-HIV consortium and the HPTN 071 (PopART) study team

## Abstract

While much progress has been made in reducing the incidence of HIV-1 infection in sub-Saharan Africa in recent years, bringing the epidemic to an end will require identification of the demographic groups that continue to contribute to transmission. Pathogen phylogenetics and individual-based mathematical models (IBMs) of transmission are approaches that enable researchers to explore such questions. Here, we used both methods to characterise the ages and sexes of the individuals involved in heterosexual transmission in the context of the HPTN 071 (PopART) trial in Zambia.

The two methods produced largely concordant results, strengthening confidence in both. A principal finding was that when the age gap in transmission (the difference of ages between the two individuals) was stratified by recipient age, the largest differences were for the youngest female recipients and the smallest for the youngest males. For women under 21 this stood at a male 9.87 years older (95% CI: 8.02 - 11.59) in the phylogenetics, compared to 6.93 (95% HDI: 6.56 - 7.32) in the IBM. As the age of female recipients increased, this gap decreased towards parity. Conversely, the under-21 male recipients saw the smallest gaps with the female older by 0.14 years (95% CI: −2.95 - 3.23) in the phylogenetics and 1.38 years (95% HDI: 0.98 - 1.68) in the IBM. As the age of male recipients decreased, this gap steadily increased. The consequence of this pattern is that transmission to new age cohorts first entering into sexual activity is driven predominantly by male-to-female transmission.

We also showed that targeting interventions at younger adults captures most of the benefit of population-wide approaches. We used the IBM to simulate the PopART universal testing and treatment intervention into the future, showing that effective treatment of under-35-year-olds would account for 94.3% (95% HDI: 65.8% - 126.6%) of the reduction in incidence by 2039 that would be achieved by treating the entire population, while effective treatment of under-35 men accounts for 60% (95% HDI: 23.2% - 92.1%).

Finally, we simulated a one-year cessation of ART treatment for the whole population, which resulted in an immediate increase in both incidence and the average age at transmission of both sources and recipients. The magnitude of this was 4.6 years (95% HDI: 2.17 - 6.24) for female recipients, 5.3 (95% HDI: 2.74 - 7.09) for male recipients, 5.24 (95% HDI: 2.78 - 6.97) for female sources, and 6.04 (95% HDI: 2.92 - 8.09) for male sources. These changes would be slow to reverse even after ART was restored.

These findings indicate that substantial reductions in HIV incidence can be achieved through intensified testing and treatment of individuals aged under 35, and in particular young men, a group that drives the infection of younger women and for whom engagement with care remains disproportionately low.

## INTRODUCTION

The widespread rollout of HIV testing, antiretroviral treatment (ART), and voluntary medical male circumcision (VMMC) in combination prevention programmes has resulted in considerable declines in both AIDS deaths and new HIV infections (1). ART is highly effective in preventing HIV transmission as well as clinical illness (2), and recent dolutegravir-containing regimens have reset the clock with regards to antiretroviral resistance, whose prevalence had recently been increasing (3).

However, questions remain concerning in which demographic groups transmissions are still occurring and how prevention interventions can best reach the groups that contribute most to new infections; and also whether, in the light of rapid population growth in sub-Saharan Africa (4), these groups will grow as proportions of the population.

Heterogeneities in the epidemic are well known (5–8), including variations in HIV acquisition risk by age and sex. Age and sex dynamics of heterosexual sexual relationships in SSA, regardless of HIV status, have also been explored (9–11), usually finding the male partner to be on average several (five or less) years older than the female. Less is understood about which age and sex groups are most likely to transmit HIV, and the age gap between sources and recipients of transmission.

This is partially because it is harder to identify sources of transmission than it is to identify recipients. Sources can be identified using traditional epidemiological survey methods, including recruitment of household pairs into a study (12), characterisation of the sexual contacts of recipients by contact tracing (13) or interviewing those recipients (5,14). These can be intensive endeavours, suffering from both recall (15) and social desirability (16) bias.

Identifying heterogeneities in HIV transmission by age and sex may be critical in informing the next steps of HIV prevention. For example, testing programmes typically yield fewer new diagnoses for men than for women, with consequences for the number of men linked to care. However, programmes aiming to end new heterosexual infections cannot ignore or de-emphasise the role of men in transmission, and must concern themselves with the male population. The risk of being a source of HIV infection will not be evenly distributed amongst males. If subpopulations disproportionately driving new infections can be identified, then the focus and impact of interventions could be enhanced by tailoring services to meet those men’s needs.

Survey-based research into heterogeneities in age and sex in HIV transmission in SSA has produced varied and sometimes contradictory results. A consistent observation, in many studies conducted during the last two decades, has been that a large age gap with a partner is associated with increasing HIV prevalence or acquisition risk in (usually young) women (9,17). However, some recent studies did not observe a simple increase in risk with increasing age of the male partner (5,7,14,18–20), suggesting that a new epidemic environment with widespread ART use has changed transmission dynamics (7). In this environment, partnerships most likely to result in HIV infection would be with someone who is old enough to have acquired HIV, but has yet to be diagnosed and started on ART. The effect would be to reduce the average age of sources of transmission, and in turn reduce the gap between source and recipient ages. The most recent studies, however, did find the original association (21,22). Studies of this sort in men are much less common, but those that have been conducted show a similar pattern; older studies have the risk of HIV acquisition increasing for men with older or same-age partners when compared to those with younger partners (9), but with this phenomenon disappearing or becoming less straightforward in more recent work (14,23). Regardless of recipient sex, characterisation of the demographics of sources in this study design is generally rudimentary, with even their ages often simply derived from recipient interviews.

Individual-based mathematical models (IBMs) (24–26) and pathogen phylogenetics (27,28) are alternative methods to surveys for describing characteristics of sources of transmission. IBMs model entire transmission networks amongst simulated individuals with the characteristics of interest. In pathogen genomics, recent methodological developments have shown that “deep sequence” analysis of within-host viral genetic diversity can identify pairs of individuals whose HIV infections likely represent direct transmission, as well as the direction of that transmission (29–31). For example the method *phyloscanner* (*27*), validated using data on known sexual partnerships (32–34), has been used to investigate transmission patterns through the identification of transmission pairs.

A previous IBM study (35) predicted that increasing ART rollout would, due to reduced force of infection, have an “ageing” effect on the epidemic, with the average age of both sources and recipients of transmission expected to increase over time. Amongst phylogenetic studies, de Oliveira et al (6) suggested a cycle of transmission in South Africa in which women under 25 are infected by markedly older men, while female-to-male transmission occurred in older individuals of similar ages.

Significantly, they did not have access to a method to reconstruct the direction of transmission using individual-level data, and instead assumed that it was determined by the prevalence gradient in the relevant communities (36). A large age gap (a mean of 9.6 years) was also observed in heterosexual transmission in Switzerland (37). More recently Magosi et al. (8) used more sophisticated methods to infer direction of transmission (38) and did not find a similar asymmetry in age differences in Botswana. Instead, they found the age gap in transmission pairs to be small, albeit with large uncertainty. Monod et al (39), working with data from the Rakai Community Cohort Study in Uganda, identified a significant upward shift in the age of both partners in male-to-female transmissions between 2003 and 2018, leading to a declining proportion of new infections occurring amongst women aged 15-24 and an increasing one amongst those aged 25-34.

The early months of 2025 saw a sudden suspension of the President’s Emergency Plan for AIDS Relief (PEPFAR). The programme was responsible for 49% of international spending, and 34% of all spending, on HIV/AIDS in SSA during 2024 (40). While funding has not entirely ended, as of the time of writing its future still remains highly uncertain (https://www.nytimes.com/2025/07/23/health/pepfar-shutdown.html). Any suspension, even if temporary, of such a crucial component of existing HIV care in SSA would likely have a profound impact on health outcomes in the region. ART prevents both progression to AIDS and onward transmission of the virus. As a result, a cessation or pause in viral suppression for the millions of HIV-positive individuals who have relied on treatment provided under PEPFAR would result in not only a significant increase in the number of individuals progressing to AIDS, but also in incidence of new HIV infections. Modelling studies have predicted that:

● In Zimbabwe persistent cuts would lead to a 78% increase in HIV incidence in Zimbabwe by 2030 (41),
● In seven sub-Saharan African countries, a 90-day funding freeze would result in between 35,000 and 103,000 new infections, depending on modelling scenario (42)
● In low and middle income countries globally, cuts in aid spending coupled with the withdrawal of PEPFAR would cause between 4,43 and 10.75 million deaths between 2025 and 2026 (43).

There is an urgent need to understand the epidemiology of HIV in SSA as we move into a new era of reduced international funding of HIV programmes (44).

Here, we provide a comprehensive characterisation of the age- and sex-specific transmission dynamics during the large HPTN 071 (PopART) trial in Zambia using two independent methods: an IBM and pathogen genomics. We use these to investigate the potential impact of future interventions focussed on suppressing transmissions from inferred source populations. We conclude that there is a substantial opportunity to achieve substantial reductions in HIV incidence by improving testing, treatment, and ART adherence of men aged 25-34. We also investigate the effect of an immediate pause of one year of ART provision on these dynamics, finding that it would cause a sudden and dramatic increase in the age of infection for both sexes.

## RESULTS

### Data

HPTN 071 (PopART) was a cluster-randomised trial of a combination HIV prevention package involving universal testing and treatment in 21 communities, comprising one million individuals, in Zambia and South Africa. It was conducted between 2014 and 2018 (45,46). The intervention involved annual household visits from community health workers offering HIV counselling and rapid testing, followed by linkage to care for HIV-positive clients. The trial had three arms, with arms A and B testing variations of the package and arm C serving as a control arm; for full details see the trial paper (46). To estimate intervention impact, HIV incidence was measured in a cohort of around 2,000 randomly selected individuals in each community, called the Population Cohort (PC). The IBM ‘PopART-IBM’ was developed as part of the trial to simulate the epidemic and interventions to help understand trial findings and extrapolate them to different settings and timescales (47,48). The IBM was calibrated to various sources of age- and sex-stratified data from the trial. In simulating the whole epidemic, the IBM generates a large number of transmission pairs on a timescale spanning the entire HIV pandemic; for comparability with the phylogenetically identified pairs (described next), from these we selected for analysis a subset which would be eligible to be sampled as part of the phylogenetic study. Across the 1000 replicates of the simulation (see Methods) there were a mean of 213,542 transmissions with recipients in this set, with a 95% highest density interval (HDI) of (191,652 - 231,239).

An ancillary study to the main trial—the HPTN 071-02 (PopART) Phylogenetics Study—was conducted as part of PopART in the Zambian PopART communities only. The study used phylogenetics to identify and characterise sources of HIV transmission in these communities, thus informing modelling of the impact of different intervention scenarios. For this purpose, viral genomic deep-sequencing was performed on samples collected over the course of the trial from both HIV-positive individuals in the PC and HIV-positive individuals attending health-care facilities in the study communities who were not on ART at the time of attendance.

While enrollment in the former was capped at 45 years of age, there was no upper age limit for inclusion in the latter. Analysing the resulting sequence data with *phyloscanner*, we identified 355 opposite-sex transmission pairs with high confidence in the direction of transmission.

The transmission pairs identified phylogenetically and those simulated from the IBM give us two independent means of characterising transmission during the trial.

### Two independent methods agree on sex and age differences in transmission

Male-to-female transmissions made up 56% (198/355) of the phylogenetically identified pairs and 57.9% of the IBM pairs (95% HDI: 56.3% – 59.7%). This is in contrast to males making up, at the start of 2017 in the IBM, 41.6% (95% HDI: 39.9% - 43.3%) of infected individuals and 44% (95% HDI: 42.2% - 45.8%) of virally unsuppressed individuals. To estimate the age at transmission of both partners in each phylogenetic pair, we used an estimate for the time of infection of the recipient derived from the HIV-phyloTSI method (49), together with the dates of birth of both partners. The age, sex, and other characteristics of the phylogenetic pairs are summarised in table 1. Figure 1 shows the distribution of source and recipient ages at the time of transmission, by sex, as estimated by both the IBM and the phylogenetic data. Table 2 shows the mean ages, by sex and direction of transmission in the pairs identified using both methods. Note that the difference in methodologies dictates that the phylogenetics estimate is given as a frequentist mean and confidence interval (CI), while the IBM is a posterior mean of means (PMM), together with the 95% HDI for the mean. The uncertainty intervals are not directly comparable. A posterior predictive check of the phylogenetic means against the posterior indicates evidence against the phylogenetics sample coming from the IBM posterior for the female-to-male age gap (see the last column of table 2).

**Figure 1:**
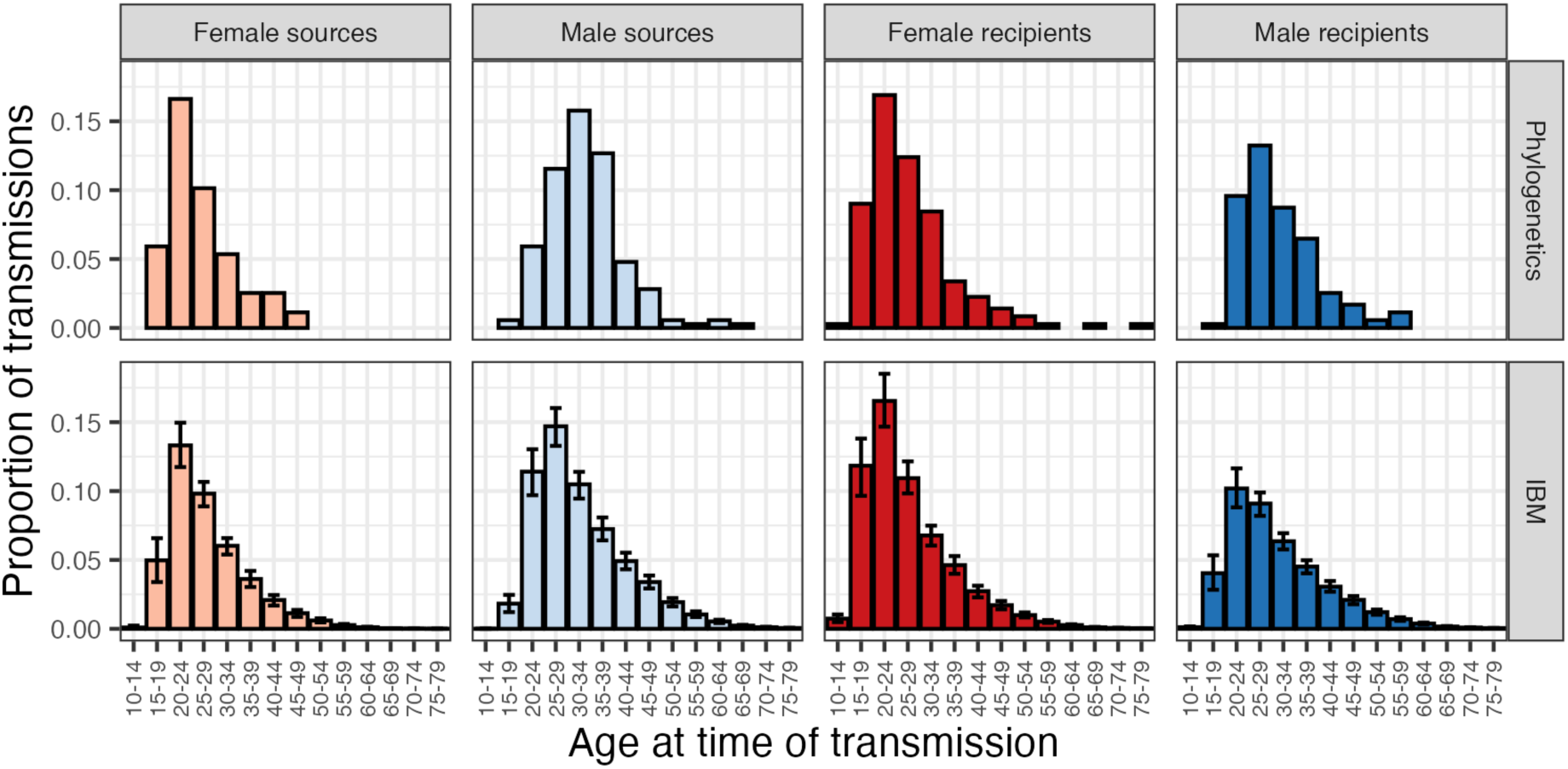
Phylogenetic analysis and the IBM model show similar age distributions for sources and recipients. Histograms of the proportions of transmissions from and to different age groups in the IBM results (top) and the phylogenetics (bottom). Values from the IBM are the posterior mean proportion of transmissions with the 95% highest density interval displayed. As every transmission has a source and a recipient, proportions sum to 1 across the two source histograms (left, light colours) and also to 1 across the two recipient histograms (right, dark colours).

**Table 1:**
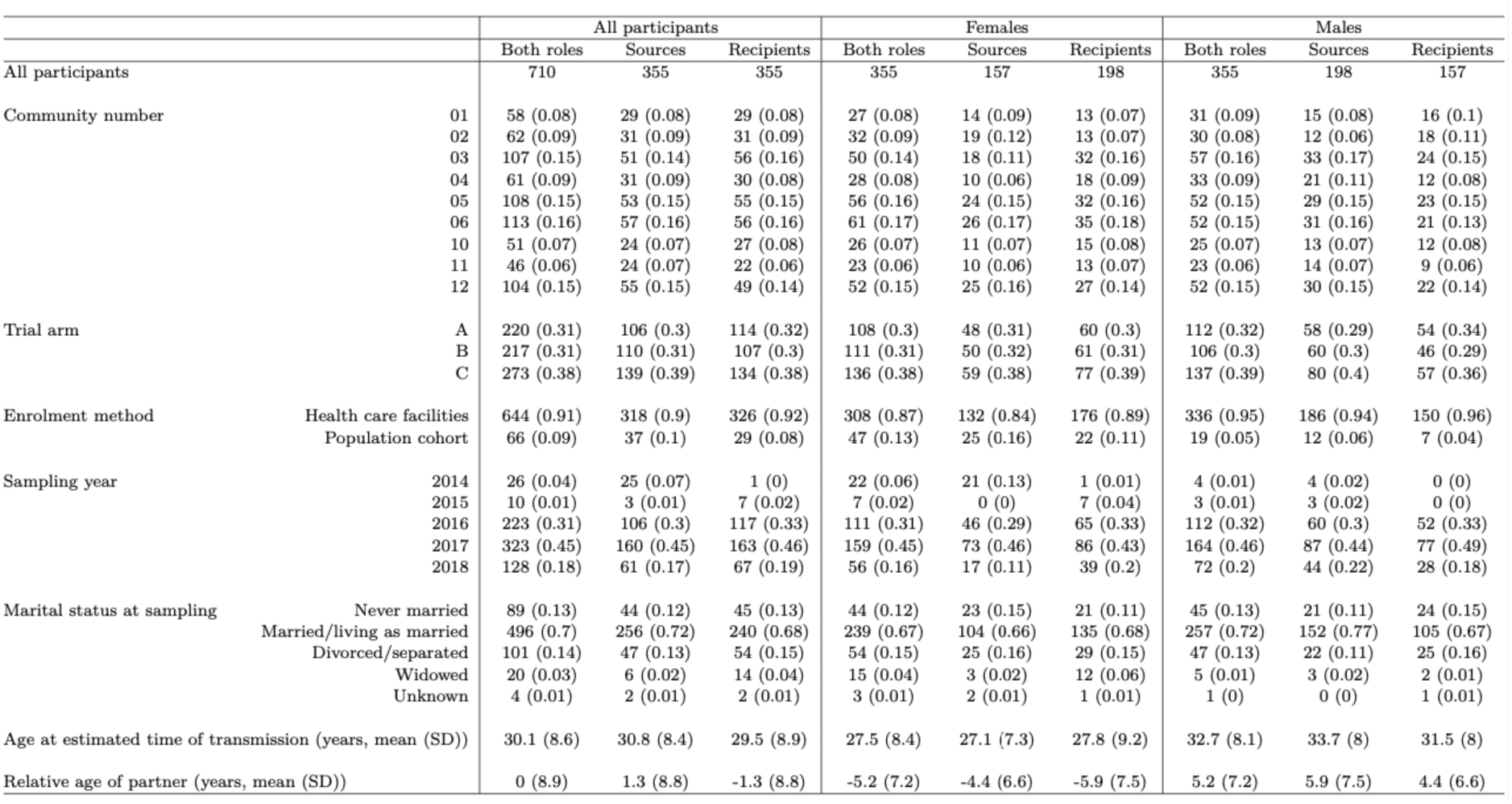
Demographics of the individuals in the 355 probable transmission pairs. Age is calculated at the estimated time at which the source infected the recipient. Numbers are given as count and proportion (column-wise) for discrete variables and mean and standard deviation for continuous variables.

**Table 2:** Summary of ages, and age gaps, in transmission pairs identified using both methods. The posterior predictive *p*-value is for the null hypothesis of the mean age or age gap from the phylogenetics coming from the posterior distribution of means from the IBM.

| Variable | Sexes | Phylogenetics |  | IBM |  | Posterior predictive p-value (phylo vs IBM) |
| --- | --- | --- | --- | --- | --- | --- |
|  |  | Mean | 95% CI | Posterior mean of means | 95% HDI |  |
| Source age | Males | 33.7 | (32.6 - 34.8) | 32.8 | (31.9 - 33.6) | 0.21 |
|  | Females | 27.1 | (25.9 - 28.2) | 28.3 | (27.4 - 29.2) | 0.11 |
| Recipient age | Males | 31.5 | (30.3 - 32.8) | 31.1 | (30 - 32.1) | 0.61 |
|  | Females | 27.8 | (26.5 - 29.1) | 27.5 | (26.7 - 28.4) | 0.71 |
| Age gap (source-recipient) | Male source, female recipient | 5.87 | (4.82 - 6.92) | 5.27 | (4.98 - 5.57) | 0.27 |
|  | Female source, male recipient | -4.44 | (-5.49 - -3.4) | -2.83 | (-3.17 - -2.47) | 0.002 |

Estimates of the distribution of transmission age gaps (source minus recipient) are shown in figure 2 and summarised in table 2. For phylogenetics there was weak evidence of a difference in the gap separating the partners’ ages between male-to-female and female-to-male pairs (*t*-test *p*=0.058). In the IBM, the gap was 2.44 years (PMM; 95% HDI: 2.11 - 2.83). A more detailed illustration of the transmission dynamics can be seen as a who-aquires-infection-from-whom (WAIFW) matrix in figure 3.

**Figure 2:**
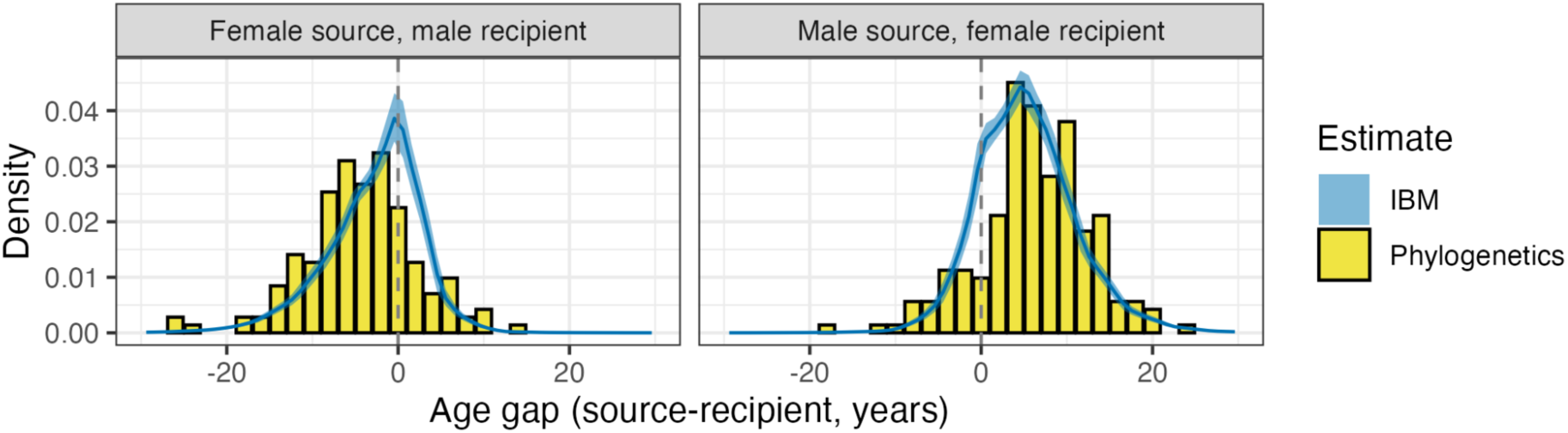
Male partners are on average older in both male-to-female and female-to-male pairs. Distributions of age gaps within transmission pairs from the phylogenetics (yellow, histogram) and the IBM (blue, kernel density estimate). For the IBM, the 95% highest posterior density intervals are given by the shaded area. Female-to-male transmission pairs are displayed on the left and male-to-female on the right. The dashed line indicates equal age.

**Figure 3:**
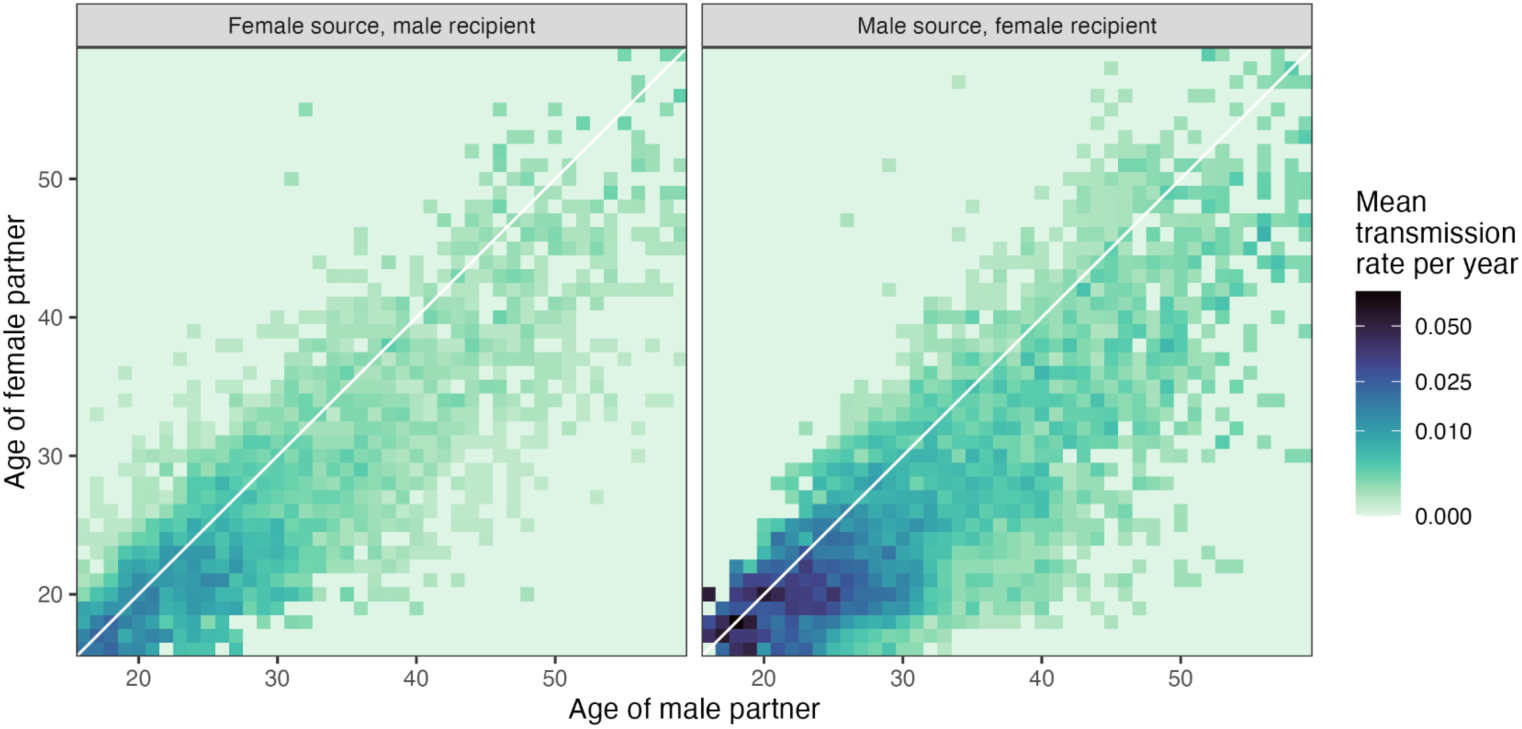
Transmissions are most common in young couples. Age-based who-acquires-infection-from-whom (WAIFW) matrix for modelled transmissions occurring in 2016. Values are the mean, over the entire year, of the per-year transmission rates from infected individuals in one age group to individuals in another (given by the number of transmissions divided by the size of the source population).

### The age gap increases with recipient age in female-to-male pairs and decreases in male-to-female pairs

The IBM provides a full transmission tree from which the complete dynamics of transmission can be explored; as inclusion in the dataset is automatic, biased sampling is not an issue. With the phylogenetic results providing independent support for its accuracy, we proceeded to explore its outputs in more detail. Figure 4 shows alluvial plots for the transmission history of individuals who became infected with HIV in the year 2016 in the best-fitting IBM simulation. Orange and green flows show viral flow to younger and older groups respectively. For female-to-male transmissions we see that these flows are almost equal, whereas male-to-female transmission are overwhelmingly orange, i.e. to younger ages. Thus this shows that there is a clear shift in age for male-to-female transmissions which is absent for female-to-male transmissions, acting to “rejuvenate” the epidemic.

**Figure 4:**
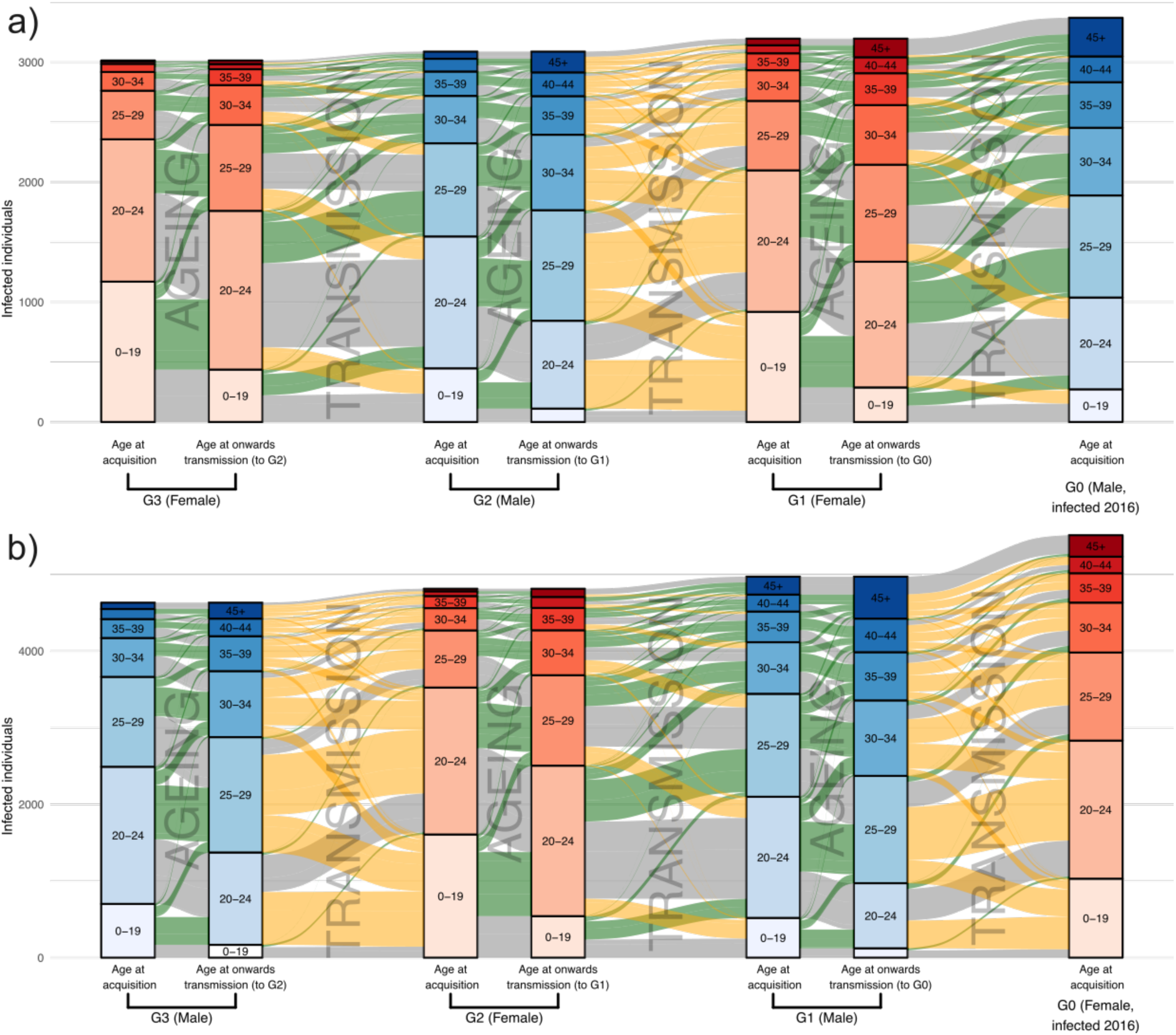
Transmissions to a younger cohort take place predominantly in male-to-female transmissions. Alluvial plots for the transmission history, derived from the IBM, of a cohort of males (panel a) and females (panel b) infected with HIV in 2016. That cohort is G0, shown on the right; to the left of it are the three previous generations of transmission (indicated as G1 to G3) that gave rise to the infections of G0. Male generations are blue and female red. The partitioning within each vertical bar indicates its age distribution. The age distributions of G1 to G3 are shown at both the time of their own infection (left) and the time at which they infected an individual in a subsequent generation (right). Movement to an older age group (by ageing or transmission) is shown by a green alluvium, to a younger group by an orange alluvium, and to the same age group by a grey alluvium. If an individual was responsible for two or more infections in the subsequent generation, then they still contribute a total of 1 individual to their “age at onwards transmission” bar, which is split proportionally between age bands depending on the number of infections they were responsible for while they were a member of each band. (For example, if individual A infected two individuals while they (A) were aged 20–24 and one individual while they were aged 25–29, A would contribute ⅔ of an individual to the 20–24 bar and ⅓ to the 25–29 bar.) Note that the members of G0 are all members of a particular cohort who were infected with HIV, but all other generations are composed solely of people who transmit HIV, a somewhat different group.

As this finding seemed contrary to the fairly consistent central estimates of the age gap described above, we stratified the gap by recipient age (figure 5a,b). This showed distinctly different patterns for the two directions. In female-to-male transmission pairs the average age gap for the youngest recipients actually had the female partner older, but this rapidly reverses as recipient age increases, with the gap decreasing in a roughly linear fashion. For male-to-female pairs there is also a decline in the gap with increasing recipient age, but the decline levels out in a fashion appearing to be non-linear. These trends are similar in the phylogenetics data (figure 5c,d), although the magnitude of the age gap for the youngest female recipients is larger, with the mean gap for recipient women aged under 21 years being 9.87 years (95% CI: 8.02 - 11.59), compared to a PMM of 6.93 years (95% HDI: 6.56 - 7.32) in the IBM. The equivalent numbers for young men are a female partner 0.14 years (95% CI: −2.95 - 3.23) older from the phylogenetics and 1.38 years (95% HDI: 0.98-1.68) from the IBM.

**Figure 5:**
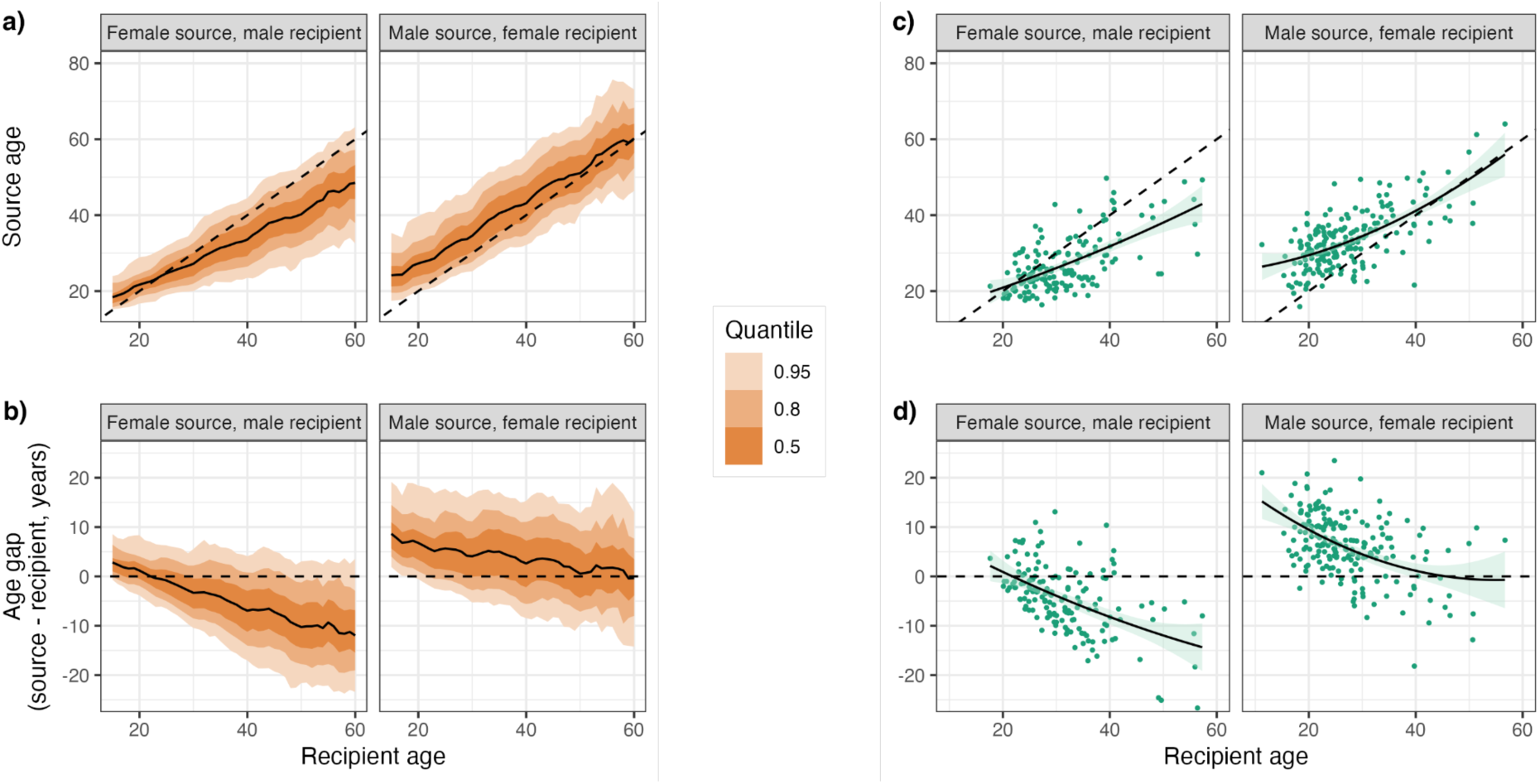
The age gap increases with recipient age in female-to-male pairs and decreases in male-to-female pairs. Relationship between recipient age and source age (a, c) and age gap (b, d) from the IBM (a, b) and the phylogenetics (c, d). The dashed line represents equal ages. The IBM results are from the “best-fitting” simulation (that with the minimum ABC distance) and represent the mean value (solid black line) and quantiles (orange shaded areas) of the y-axis variable in 1-year increments. The individual values from the phylogenetics pairs are plotted as green points, with a regression line fitted using quadratic polynomial regression and the 95% confidence region given by the pale green area.

### Transmissions from an undiagnosed source involve younger individuals

Using the IBM, we stratified the age and sex variables according to the diagnostic status of the source. A posterior mean of 73.8% (95% HDI: 71%-76.3%) had an undiagnosed source. This was 70.7% (95% HDI: 67.2%-73.6%) of female sources and 76% (95% HDI: 73%-78.8%) of males. Figure 6 shows variation in this proportion by age, with the largest proportions occurring in the youngest age groups for both sexes.

**Figure 6:**
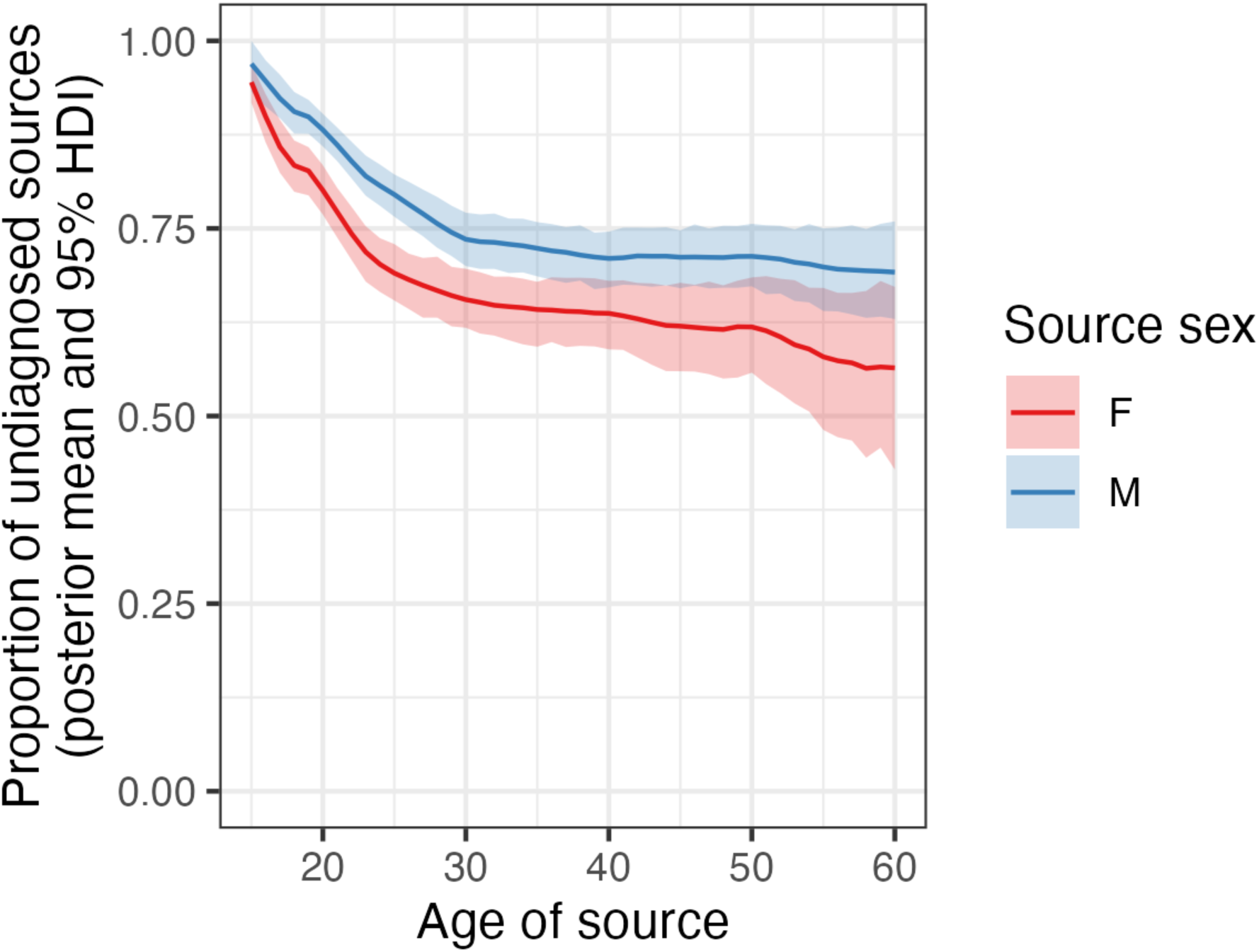
Proportion of sources that are undiagnosed, by age and sex. The thick line is the posterior mean and the shaded area the 95% HDI.

We also examined age and sex dynamics by diagnostic status (table 3). Both individuals involved in these transmissions were on average older when the source had received an HIV diagnosis, regardless of sex. Male sources, female sources, and male recipients were all a mean of around three years older when the source was diagnosed. Female recipients, however, showed a smaller increase in age, with the difference in ages between diagnosed and undiagnosed being 1.45 years (PMM; 95% HDI: 1.06-1.94). This combination meant that there was no significant difference in age gaps by source diagnostic status for female-to-male couples, with the PMM difference being 0.044 years and the 95% HDI overlapping zero (−0.12 - 0.2) but one existing for male-to-female pairs (PMM 1.09, 95% HDI 0.9 - 1.27).

**Table 3:** Summary of ages, and age gaps, in transmission pairs identified using both methods, stratified by diagnostic status of the source, and the posterior mean and 95% HDI values of the difference between the mean values (diagnosed-undiagnosed)

| Variable | Sexes | Source diagnosed |  | Source undiagnosed |  | Difference of means (diagnosed-undiagnosed) |  |
| --- | --- | --- | --- | --- | --- | --- | --- |
|  |  | Posterior mean of means | 95% HDI | Posterior mean of means | 95% HDI | Posterior mean | 95% HDI |
| Source age | Males | 34.8 | (33.8 - 35.6) | 32.2 | (31.4 - 33.1) | 2.54 | (2.14 - 2.91) |
|  | Females | 30.1 | (29.2 - 31.1) | 27.5 | (26.6 - 28.4) | 2.57 | (2.23 - 2.98) |
| Recipient age | Males | 32.9 | (31.8 - 34) | 30.3 | (29.3 - 31.4) | 2.53 | (2.07 - 2.96) |
|  | Females | 28.7 | (27.7 - 29.5) | 27.2 | (26.4 - 28.1) | 1.45 | (1.06 - 2.96) |
| Age gap (source-recipient) | Male source, female recipient | 6.1 | (5.7 - 6.49) | 5.01 | (4.73 - 5.28) | 1.09 | (0.9 - 1.27) |
|  | Female source, male recipient | -2.8 | (-3.21 - -2.43) | -2.84 | (-3.19 - -2.5) | 0.044 | (-0.12 - 0.2) |

### Age patterns remain stable over time despite steep increases in ART coverage

We next investigated whether a shift in age patterns in the IBM occurred as a result of increasing use of ART (7). Figure 7 shows posterior trends in source age, age gap, and proportion of individuals on ART over time for the period 2005–2020. For female-to-male transmissions, a small decrease was seen in both source ages and the age gap. The PMM female source age was 29 (95% HDI: 28.1 - 30) years in 2005 and 27.4 (95% HDI: 26.1 - 28.7) in 2020, while the age gap had the male 3.07 years older (PMM; 95% HDI: 2.68 - 3.46) in 2005 and 2.63 (95% HDI: 2.23 - 3.06) in 2020. In male-to-female transmissions, source ages declined from 33.5 years (PMM; 95% HDI: 32.5 - 34.3) in 2005 to 32.5 (PMM; 95% HDI: 31.5 - 33.4) in 2012, but then levelled off, with the mean being 32.5 (95% HDI: 31.3 - 33.6) in 2020. Age gaps were stable between 2005 and 2012, going from 5.26 years (PMM; 95% HDI: 4.93 - 5.59) to 5.21 (PMM; 95% HDI: 4.85 - 5.53), but then rose, reaching 5.48 (PMM; 95% HDI: 5.06 - 5.91) by 2020.

**Figure 7:**
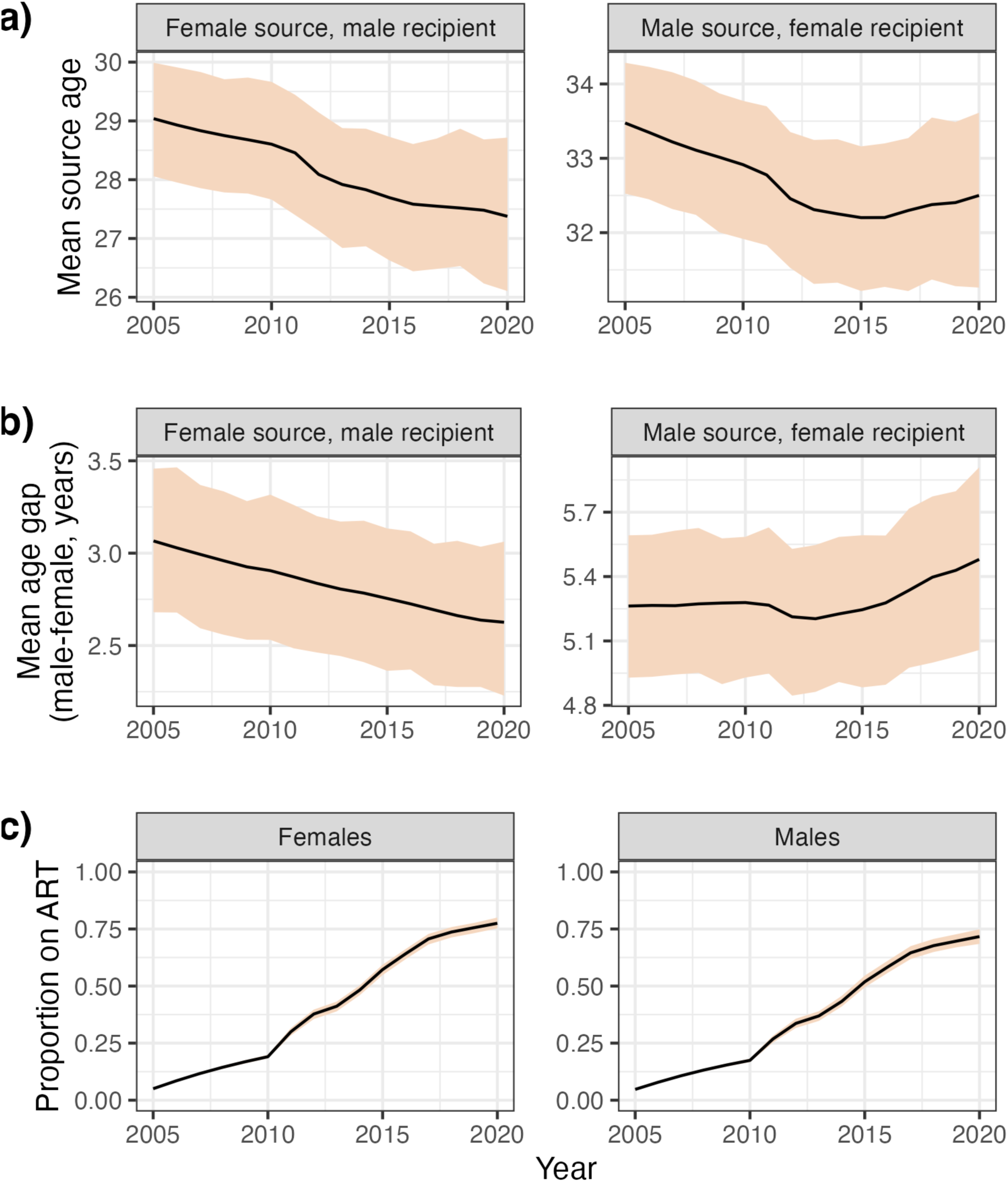
Little change in age gaps over the period of ART rollout. Time trends in age of sources (a), age gaps (b) and proportion of the population on ART (c) over time. Estimates are the posterior mean (black line), and 95% HDI (shaded), of the mean values.

### Rollout of the PopART intervention to individuals aged 35 or under is projected to reduce incidence by 67% by 2040

We used the IBM to quantify the effect of targeting future interventions at different demographic groups. We started from the model state of the entire population of the control communities (those which were not subject to any part of the trial intervention and received standard of care) at the end of 2019 (47). We considered alternative hypothetical scenarios where the PopART intervention was used as a proxy to deliver real-world intensive linkage to care programmes to specific demographic subgroups. We focus a) on younger age groups, since our data shows that they are more likely to be unsuppressed than older age groups living with HIV and b) on younger men, who were shown to drive the rejuvenation of the epidemic (figure 4).

We modelled the effect of four different scenarios, starting in the year 2020:

1. Applying the PopART intervention to the entire population
2. Applying the PopART intervention to only the population aged between 18 and 34, with standard of care for other age groups
3. Applying the PopART intervention to only the male population aged between 18 and 34, with standard of care for other male age groups and females
4. Standard of care

Figure 8 shows the effects of these four scenarios on HIV incidence, by two-year period from 2018–2019 (prior to the introduction of the intervention). The posterior mean incidence in 2018–2019 was 0.8 cases per 100 person-years at risk (95% HDI: 0.592 – 1.01). By 2038–2039, overall incidence was projected to decline to 0.43 (95% HDI: 0.2 – 0.693) even in the absence of any PopART intervention. The PopART intervention, when provided to the entire population, further reduces this to 0.12 (95% HDI: 0.001 – 0.25). However, the effect of deploying it only to individuals aged 18-34 has an only slightly smaller effect, with a posterior mean incidence of 0.14 (95% HDI: 0.024 – 0.27), while providing it only to men aged 18-34 gave an incidence of 0.24 (95% HDI: 0.067 – 0.42). By the end of 2039, the former intervention achieved 94.3% (95% HDI: 65.8% - 126.6%) of the reduction between the “standard of care” scenario and full deployment (the stochastic nature of the IBM meaning that the 18-34 intervention could outperform the full intervention in some cases), and the latter 60.0% (95% HDI: 23.2% - 92.1%). The 5.78% (95% HDI: 4.7%-7.1%) of the population that remained virally unsuppressed in 2018-19 would be reduced to 0.38% (95% HDI: 0.072% - 0.76%) by 2038-39 under the full intervention, to 0.62% (95% HDI: 0.023% - 1.04%) when deployed only to those aged 18-34 and and to 1.17% (95% HDI: 0.45% - 1.85%) when deployed only to males 18-34.

**Figure 8:**
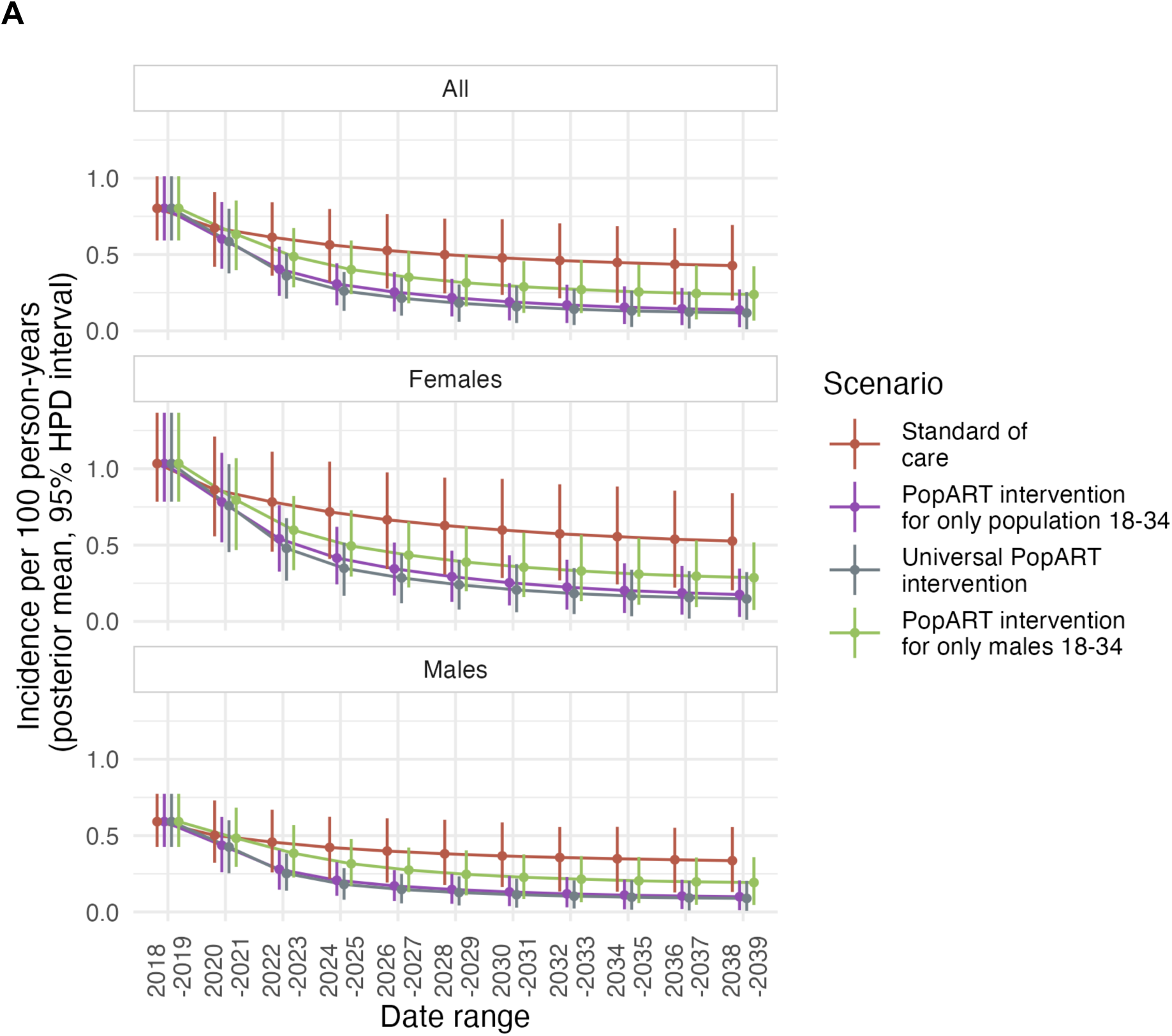
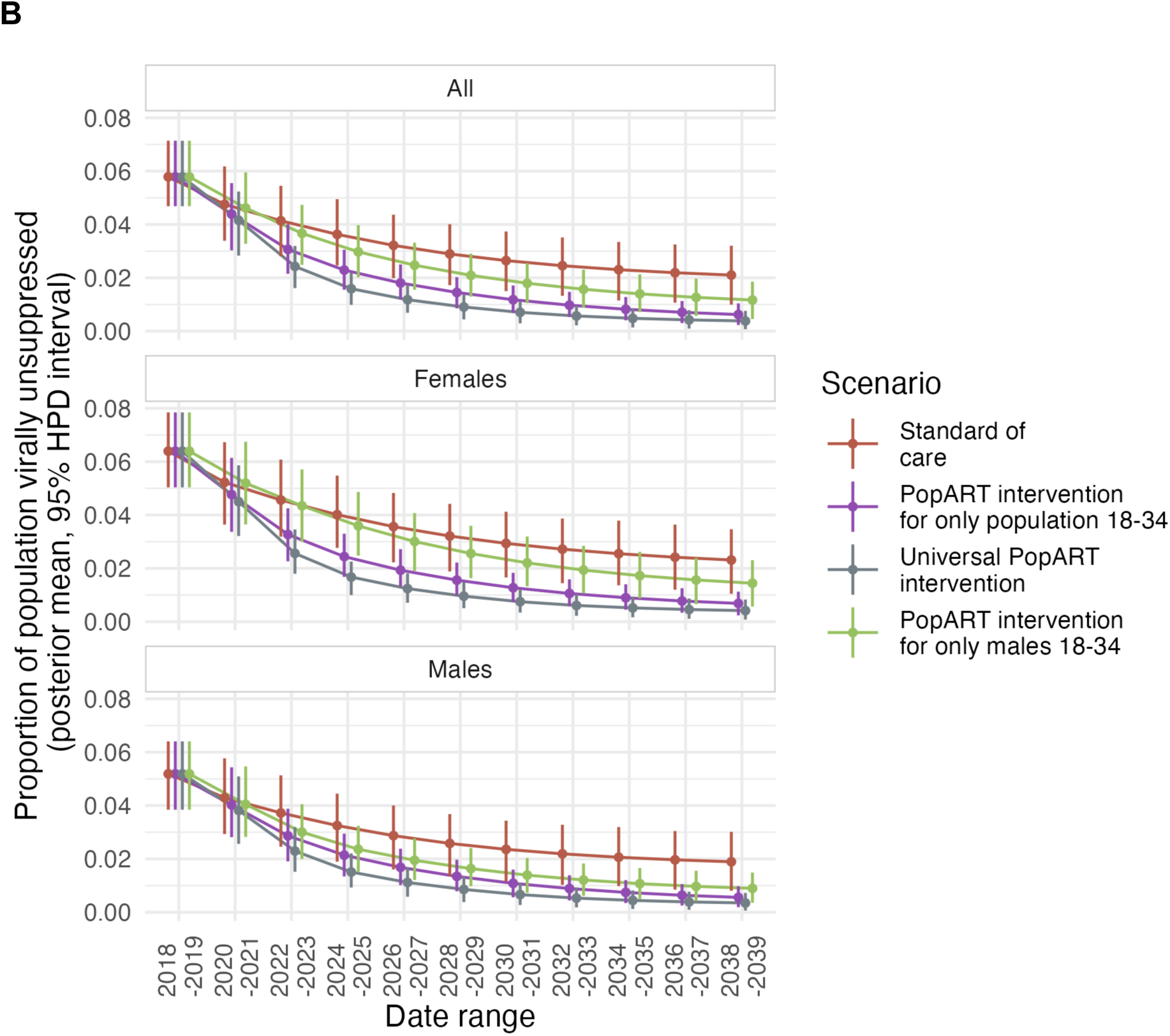
The impact of an intervention focussing on younger people. Projected time trends in HIV incidence (A) and population proportion of people with unsuppressed HIV (B), 2018–2041, in the Zambian control communities of the PopART trial, under various scenarios of future PopART intervention rollout. Posterior mean estimates are calculated in two year time periods for the whole population (top), females (middle) and males (bottom). The four scenarios are, firstly, no CHiPS deployment at all (red), provision of the full PopART intervention to the whole population (purple), PopART intervention targeting only those aged under 35 (grey), and PopART intervention targeting only men aged under 35 (green). Vertical bars indicate the 95% highest posterior density interval.

### Modelling a one-year gap in ART provision

Finally, again starting with the model state in the control communities, we modelled a one-year complete pause in ART provision for the year 2025. Following this, re-engagement with care took place following a subsequent positive test; no deployment of additional resources or targeted interventions to re-integrate individuals with care was modelled. We examined the effect of this cessation in ART on age and sex dynamics, which can be seen in figure 9. The pause is responsible for a substantial increase in incidence (fig 9a), with a posterior mean of 2067 (95% HDI: 817 - 6708) new cases in 2024 increasing to 6086 (95% HDI: 3463 - 10172) in 2025. Regarding age and sex dynamics (figure 9b), the immediate effect was an increase in the ages of all those involved in HIV transmission. The change in the ages of all individuals involved in transmissions from 2024 to 2025 was 4.6 years (PMM; 95% HDI: 2.17 - 6.24) for female recipients, 5.3 (PMM; 95% HDI: 2.74 - 7.09) for male recipients, 5.24 (PMM; 95% HDI: 2.78 - 6.97) for female sources, and 6.04 (PMM; 95% HDI: 2.92 - 8.09) years for male sources. There was also a jump in the age gap in male-to-female transmissions, going from a gap of 5.34 years (PMM; 95% HDI: 4.38 - 6.28) in 2024 to 6.78 (PMM; 95% HDI: 5.73 - 7.96) in 2025. No similar effect was seen in female-to-male pairs. After the initial change in 2025, the curves in figure 9 can be seen slowly returning to the patterns of the years 2019-2024, but in the absence of specific strategies to re-engage the HIV-infected population with care, this is not immediate once ART provision is re-established.

**Figure 9:**
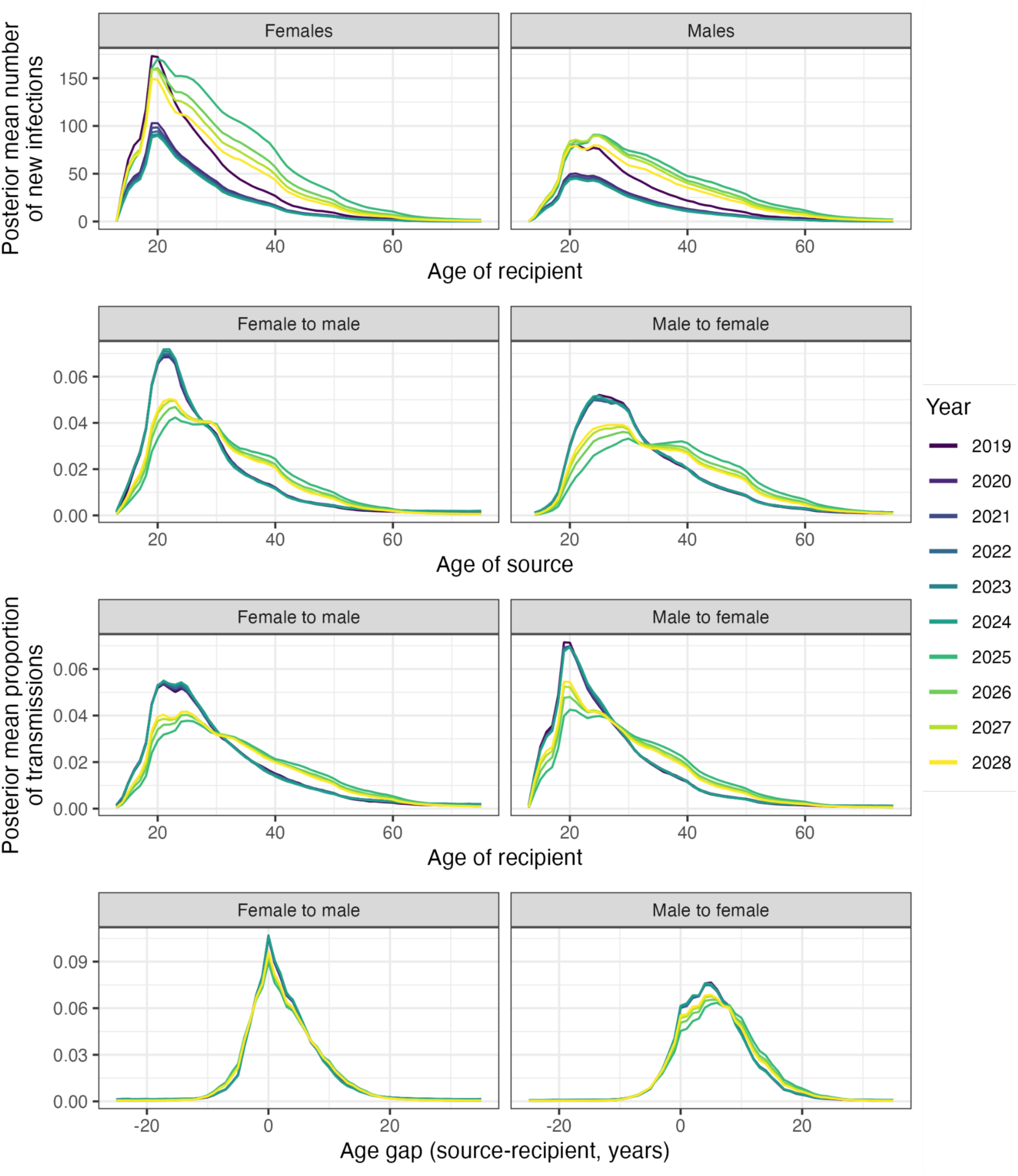
The impact of ART interruption. A: Posterior mean new infections by year, by sex and age, and B: the age distribution of new transmissions by year, in the circumstances of a complete cessation of ART provision during 2025, returning in 2026. Lines chart the posterior mean proportion of new infections for sources and recipients of each integer age (top and middle), and age gap rounded to the nearest integer (bottom). In both cases, lines are coloured by year.

### Sensitivity analyses

Appendices 1 and 2 describe two supplemental analyses of the phylogenetics data. First we confirmed that the analysis was genuinely informative, differing from results obtained by choosing pairs of individuals at random as “transmission pairs”. Second we performed a detailed sensitivity analysis of the effects of varying the settings for the *phyloscanner* package. While the sets of transmission pairs obtained using different settings were somewhat variable, the exact choice of pair set had little effect on the main results of this study.

## DISCUSSION

In this study we investigated the age and sex patterns of heterosexual HIV transmission in Zambia during the HPTN071 (PopART) trial between 2014 and 2018, using both an individual-based mathematical model (IBM) of the epidemic and pathogen phylogenetics. We focussed on characterising transmission pairs, jointly analysing sources and the recipients to whom these individuals transmit.

The population of sources — the subset of people living with HIV who transmit onwards — has been characterised in several previous studies (8,39). For the HPTN071 (PopART) trial we previously reported that male sources were on average a few years older than female sources, that there was no disproportionately large contribution to transmission from external communities or from individuals with drug-resistant viruses, and that there was a disproportionately large contribution to transmission from men aged 25–34 (50). These men should be a focus of efforts to prevent onward transmission. Interventions are needed to find them, link and retain them in care, and encourage viral suppression, even if they are more difficult and costly to reach. In that study we found that linking one man in this age group to care is likely to have a similar effect on incidence as linking two women.

Here, we further developed previous results by characterising transmission pairs, i.e. analysing sources and their recipients jointly, not independently. Notably we estimated the distribution of the age gap within a pair, separately for male-to-female and female-to-male transmission. Most transmissions in either direction had the male partner a few years older than the female, with means of between 2.83 and 5.87 years depending on data source and direction. Gaps of these magnitudes are consistent with reports of partnerships in the general population (regardless of HIV status) in the region (14,51–53).

We also observed more complex dynamics (figures 4 and 5), once recipients wre stratified by age. For male recipients, at the youngest ages the age gap is near parity, with the female source being a posterior mean of 1.38 (95% HDI: 0.98 - 1.68) years older for males under 21 in the IBM. These men would tend to take a partner younger than themselves (judging by the patterns in older men). However, such partners, if sexually active at all, are unlikely to be HIV-positive because there is a delay, which we here refer to as the “exposure lag”, between the expected age of first potential exposure to HIV and the expected age of infection. As the age of male recipients increases, the age gap reverses in direction and then increases.

For female recipients, at the youngest ages the gap is particularly large, with the male partner being a posterior mean of 6.93 (95% HDI 6.56 - 7.32) years older for women under 21. The exposure lag means that sexual partners who are HIV-positive are older than the average partner, and in addition the contribution from similarly-aged and younger men is reduced as few of those have reached sexual debut. As the age of female recipients increases, this initially large age gap decreases steadily until it reaches parity for recipients in their early 60s. Women form new partnerships with men on average a few years older than themselves, and this remains true throughout their life (11). Thus this decline in the age gap in HIV transmission pairs is likely to be driven by a combination of ART treatment and AIDS-related mortality in older men. For middle-aged and older women, the population of potential male partners will thus be increasingly virally suppressed or HIV-negative. It should be emphasised that, while HIV prevalence increases with age, only a relatively small proportion of new infections occur in older age groups and the contribution of these to the epidemic is relatively minor; across the IBM a posterior mean of only 4.66% of transmissions (95% HDI: 3.85% - 5.45%) were from individuals aged 50 or greater.

The consequence of this very divergent pattern of age gaps amongst the youngest male and female recipients is that male-to-female transmission is primarily responsible for the infection of new age cohorts as they reach sexual debut. This can be most clearly seen in figure 4, where the youngest female recipients (pale pink) are the recipients of large flows from older male groups (orange flows) while males of the same ages (pale blue) have smaller incidence, with a much greater proportion of that incidence coming from sources of the same age group.

A previous paper published by de Oliveira and colleagues (6) interpreted transmission patterns in SSA as a cycle in which young women are infected by much older men (the “sugar daddy” phenomenon), then these women age and later transmit back to similarly-aged men. The study was based on phylogenetic evidence but did not use genomic data to reconstruct the direction of transmission. Our results concur that the widest age gap in transmission to females occurs in the youngest recipient age group. Our IBM estimate has male sources a posterior mean of means 6.29 years (95% HDI: 6.01 - 6.62) older than female recipients when the latter are under 25, compared to the mean gap of 8.7 years (95% CI: 6.8 - 10.6) estimated by de Oliveira et al. In our IBM the under-25 female age group makes up a posterior mean of 50.3% (95% HDI: 45.9% - 55.2%) of female recipients, and 29.1% (95% HDI: 26.7% - 32.4%) of all recipients, indicating that this route plays a very substantial but not overwhelming role in overall transmission.

Our results diverge from those of de Oliveira et al. when it comes to female-to-male transmission, as we see the smallest age gaps for the youngest male recipients and a gradual increase with age. Men aged 18 were infected by women a posterior mean of means of 1.24 years older (95% HDI: 0.76 - 1.72), but by age 25 this gap had the women 0.92 years younger (95% HDI: 0.68 - 1.19), and by age 40, 6.88 years younger (95% HDI: 6.08 - 7.64).

Another limitation of the de Oliveira et al approach was that it collected participants into large age groups rather than considering age as a continuous variable, which may mask subtle dynamics of the age gaps in transmission, an observation also made by others (33,54).

At the population level, we see that young women are infected by men who are on average up to a decade older (depending on method). These men will be in their mid-20s to early 30s, and we do not find evidence of a major driver of new infections from men considerably older than that. At the individual level, however, it will remain true that having a much older partner poses an increased risk of HIV acquisition for a woman under 25, as often seen in previous work (21,22). An older partner has had more time to be exposed to HIV themselves, as evidenced by the phenomenon also existing for new infections in men (23).

Of the other phylogenetic studies of these questions, Magosi et al (8), in Botswana, found male partners to be a mean of 3.8 years older in male-to-female transmission pairs (with a standard deviation of 6.9 years) and female partners a mean of 1.3 years older in female-to-male pairs (with a standard deviation of 12.7 years). These large standard deviations give very wide confidence intervals, but the pattern we observed of larger (male minus female) gaps in male-to-female transmissions was present. Monod et al. (39), in Uganda, identified a clear bimodal distribution in male source ages - one peak in the late 20s and another in the late 30s - which we did not find in Zambia. They did not present comprehensive estimates of age gaps. Kusejko et al. (37), in Switzerland, found a wide gap of 9.6 in heterosexual pairs (amongst those who do not inject drugs). They used a simple phylogenetic distance criterion to identify linked individuals (the paper is in part an exploration of the limitations of this approach under different distance thresholds and sampling regimes) and did not attempt to reconstruct by direction of transmission. The varied settings of these studies may also account for some or all of the discrepancies with our work.

Increasing rollout of ART between 2005 and 2020 did coincide with changing patterns of transmission (figure 7), and the suggestion in previous work that age gaps in male-to-female transmission would decrease with ART rollout (because older males were increasingly virally suppressed) gains some support. However, the magnitudes of these changes were small, with the posterior mean of mean gaps in male-to-female transmission dropping from 3.07 (95% HDI: 2.68 - 3.46) years in 2005 only to 2.63 (95% HDI: 2.23 - 3.06) in 2020. The profound change in epidemiology suggested by some previous studies (5,7,14,18) finds little support in our analysis.

We found that both individuals involved in transmission pairs with a diagnosed source were older than those involved in pairs with an undiagnosed source. This is most likely due to the time it takes for an individual to receive a diagnosis, coupled with the age assortativity of partners. Interestingly, the increase in ages for female recipients with diagnosed partners is smaller than that for males - a posterior mean change in mean difference of 2.53 (95% HDI: 2.07 - 2.96) years for males and 1.45 (95% HDI: 1.06 - 2.96) for females - which in turn affects age gaps (see table 3). The reasons for this are hard to entirely unpick but it may be driven by differences in ART uptake between men and women.

We used two entirely independent methods, each with its advantages and disadvantages. The IBM allows us to simulate very large numbers of complete transmission chains, for which we can analyse all individuals with no sampling bias or erroneous identification of pairs. However, the results are dependent on model assumptions (for example, the age mixing matrix (47)), and the individuals involved are simulated. The phylogenetics data are the reverse: there are no model assumptions and the data come from genuine cases of HIV infection, but the sample size, although large compared to similar studies, is several orders of magnitude smaller than the actual population, and inference may be affected by sampling bias and imperfect pair ascertainment. The results of the two methods being largely consistent is a key strength of this study: it gives confidence in both methods, and in additional estimates generated by the IBM for scenarios and periods for which no phylogenetic data exist. Nevertheless, some issues may affect both methods.

Notably, neither of our methods attempt to quantify the role of men who have sex with men (MSM) in transmission. They are not modelled in the IBM, and, as we analysed opposite-sex pairs only in the phylogenetic study, we do not consider their role there either. In addition, while the IBM includes heterogeneity in sexual risk-taking behaviour it does not explicitly model key heterosexual populations, such as female sex workers (FSW). Membership of such groups is unknown for participants included in the phylogenetic study, and they might also be undersampled. Of note, FSW and MSM are both groups which were not modelled in the IBM and were likely undersampled in the phylogenetic study. Our estimates of the effect of interventions (figure 8) rely on these groups not contributing significantly to transmission. We previously estimated that sex between men makes little contribution to transmission within the population studied here (50).

The posterior predictive tests of table 2, which give *p*-values for the central estimates in the phylogenetics sample coming from the IBM, provide, in the case of the age gap in female-to-male pairs, strong evidence for a difference in the estimates derived from the two methods. However, the actual magnitude of the difference in means is fairly small (1.61 years), and this would not change the interpretation of the findings. Exploring this further, while the decline in the age gap as a female recipient ages is seen in both methodologies, the magnitude of the gap for the youngest women is the main area in which the two methods differ (figure 5), with larger gaps seen in the phylogenetics. The mean gap in the best-fitting IBM for female recipients aged 20 or younger was 6.93 years (95% HDI: 6.56 - 7.32) while for the phylogenetics it was 9.87 years (95% CI: 8.02 - 11.59). It is not entirely clear why this might be. Potentially relevant limitations in the data used to fit the IBM are the absence of trial data for under-18s and the lack of care cascade information from before 2013. Alternatively, there might have been a bias towards finding older-than-average partners for younger women in the phylogenetics data. Despite this disparity, the similarity between the trends in age gaps identified with both methods is striking. The age gap between a female recipient and her male partner at the time of transmission is largest for the youngest women and declines asymptotically towards equality as the recipient’s age increases. The youngest male recipients, on the other hand, are infected by a female source of a similar age to (or even slightly older than) themselves. With increasing recipient age, this gap subsequently widens in the direction of younger source ages.

As alluded to above, a limitation of this study is the narrow geographical catchment area of the PopART trial. The results should not be assumed to hold identically in other SSA settings. We also do not control for bias in the composition of the phylogenetic dataset, both in regard to demographics and to the ability to successfully acquire a sequence from a participant providing a sample, which we assume is random. Nevertheless, the consistent results between the two methodologies should assuage fears about this, as the IBM results are not subject to sampling bias problems.

With the 95:95:95 targets being in reach in many parts of SSA, it is becoming increasingly expensive to link individuals to care using population-wide testing and messaging campaigns. There is widespread agreement that future prevention campaigns need to focus on the remaining 5:5:5 of the population. We estimated that an intensive effort to link people to care, such as the PopART intervention, would result in almost as much reduction in incidence if focussed on those aged between 18 and 34, and would retain two thirds of the benefit when focussing on men between 18 and 34 only. Thus, the key role of young men in the infection of young women reaching sexual debut, outlined in this paper, means that a considerable (although not optimal) incidence reduction can be achieved by targeting them alone. We did not methodically explore potential age cut-offs in this work, and the identification of the optimal demographic groups to target goes beyond the scope of our analysis. Nevertheless, our results show the potential benefits of focussing on groups who do contribute disproportionately to transmission. Enabling and encouraging younger men to seek and stay in care to the same extent as women would improve the health and life chances of many men, and rapidly reduce infection rates in their partners.

Monod et al (39) modelled a counterfactual scenario different from but related to ours. Where we simulated the long-term impact of an intervention based on real-world implementation data, where the hypothetical element is targeted at different demographic groups, they simulated an alternative hypothetical intervention that would either fully close the gap in viral suppression between men and women, half close it, or achieve the UNAIDS 95:95:95 treatment targets in males. Both sets of scenarios predicted that considerable benefits in reducing female incidence could be achieved by targeting a relatively small number of HIV-positive men. This undertaking would be much less intensive than targeting the much larger number of HIV-negative women at high risk of acquisition.

A major disruption in ART provision as a result of changes in the global aid landscape is now underway, with the endpoint unclear. As ART prevents both disease progression and transmission, simultaneous increases in the incidence of AIDS and of HIV infection are to be expected, and the response of the global health community will need to focus on both. In the latter case it is important to anticipate not only an increase in new infections, but in what populations they would occur. Our results suggest a shift in incidence to older age groups, caused by the withdrawal of ART from those previously virally suppressed, at which point their partners, both existing and new, are at greatly increased risk of HIV infection. This ageing of the incidence profile would linger even after the restoration of ART provision unless procedures were put into place to re-engage those individuals with care.

In summary, using two independent and largely concordant methods, we have shown that the age gap between HIV transmission partners in Zambia varies substantially with the age and sex of the recipient, and that male-to-female transmission is the primary route by which new age cohorts are infected upon reaching sexual debut. The effective treatment of men under 35 could achieve the majority of the incidence reduction delivered by population-wide interventions. At a time when the sustainability of international HIV funding is in question, understanding where transmission occurs, and where it would shift in the event of disrupted care, is essential for designing interventions that achieve the greatest impact with finite resources.

## METHODS

### HPTN071 (PopART) trial

The HPTN071 (PopART) trial has been described in detail previously (45,46). In summary, the impact of a universally delivered combination prevention package was tested in 21 communities in Zambia and South Africa, between 2013 and 2018 (45,46). The total population of the 21 study communities was approximately one million. The intervention comprised annual rounds of home-based HIV testing, including support for linkage to care and retention on ART, promotion of medical male circumcision for HIV-negative men, and additional HIV, tuberculosis (TB), and sexually-transmitted infection (STI) services. It was delivered by teams of Community HIV-care providers (CHiPs). The 21 communities were arranged in seven triplets, matched on baseline HIV prevalence, engagement with care, and geographic location (Hayes et al., 2019). Communities were randomly allocated into one of three arms within each triplet. Arm A communities received the full combination HIV prevention package and offer of immediate ART for individuals that tested HIV-positive. Arm B communities received the full combination HIV prevention package with ART initiation following national guidelines (where ART initiation was dependent upon an individual’s CD4 count). Arm C communities received the standard of care. National guidelines on ART eligibility changed over the period of the trial, and thus from late 2016, ART for all HIV positive individuals was standard care in both countries (aligning arms A and B). The primary endpoint was the relative reduction in HIV incidence between arm A or B communities and their matched arm C community between 12 to 36 months after trial start. This was measured in a cohort of 2000 randomly selected individuals aged 18-44 per community, called the Population Cohort (PC), who were followed up in four annual rounds. Rounds of the PC are referred to as the number of months after trial start (e.g. PC0 is 0 months after start).

### IBM

For each of the 21 communities in the HPTN017 (PopART) trial, the IBM (47) models every individual in a growing population that reaches a size of approximately 50,000 in 2014, roughly matching the actual community sizes at that time point.

Demographic parameters follow historical estimates of mortality and birth rates from the UN Population Division (55), and future projections of these measures follow the UN’s intermediate estimate (56). HIV infections are introduced into the model in 1980, HIV transmission is modelled in serodiscordant couples, and HIV disease progression (in the absence of ART), follows data from the ATHENA cohort in the Netherlands (57). While the disease progression data are derived from studies of subtype B, previous work has found no significant difference in CD4 progression by subtype (58,59)

Individuals are categorised into one of three groups of sexual risk-taking behaviour, where membership of a higher risk group reflects both a higher propensity to form partnerships and an increased maximum number of concurrent partnerships. An individual’s risk group is assumed not to change through time. Partnerships are formed based upon an individual’s age, sex, and sexual-risk taking behaviour, with sexual risk-taking behaviour acting independently of age and sex. Partnership formation and dissolution rates are calibrated using survey data from the PC, and incorporate assortativity by risk group; see (47) for full details. Age mixing was determined using the age of the last reported partner (within the last year) based upon extensive sexual behaviour surveys. To allow for transmissions from areas outside the trial communities, the IBM models two “patches” representing the community and the surrounding area. Partnerships may also form with individuals in the surrounding area, but at a reduced rate (45% of the rate within the community). When serodiscordant, these between-community partnerships are also at a lower (29% of within-community) risk of HIV transmission, reflecting higher rates of condom usage reported in the PC data.

Testing and linkage to care were modelled as two separate processes, a “background” component representing testing outside the context of the trial (e.g. through clinics and antenatal care) and a “CHiPs” component representing the trial intervention, which was disabled for the control communities. The subsequent care cascade following a positive test was determined by the component from which the test came; for full details see (47).

Parameters for partnership formation and dissolution were set using data from the PC. Other parameters were estimated using Approximate Bayesian Computation (ABC) to fit the model to five sources of data, all stratified by age and sex: 1) HIV prevalence from the Zambian Demographic and Health Surveys (DHS) of 2002, 2007 and 2013; 2) HIV prevalence from the final intervention round of the trial; 3) the proportion of individuals aware of their status among HIV-positive individuals from all three intervention rounds; 4) the proportion of individuals on ART among those who are HIV-positive and aware of their status from all three intervention rounds; and 5) the proportion of individuals virally suppressed amongst all HIV-positive individuals at PC24. For all but 1), which was taken from previous work, fitting used trial data from an intervention community (arm A) in Zambia. The calibration framework fits the epidemic selecting 1000 most likely parameter sets, each of which is used to generate a single simulation. An ABC distance measures distance between observed and simulated summary statistics used in the fitting process.

Point estimates for key quantities presented here are the posterior mean and 95% highest posterior density (HPD) interval over 1,000 ABC simulations. We also identified the ‘best-fitting’ IBM simulation - the model replicate whose ABC distance was smallest - and used the corresponding transmission tree when we presented results that require a single realisation of the simulated epidemic.

The IBM was used to model transmission over a 75-year period from 1975 to 2050. To make the outputs comparable to the phylogenetics data, we selected only transmissions for which the recipient would have been eligible to provide a viral sample during the trial, specifically that they were infected with HIV before 2019 and alive in November 2013. (We did not model the viral dynamics that would result in the failure of a HIV-positive individual to provide a viable sequence; effectively we assume that the probability of this happening is independent of participant demographics or time of infection.) We also used only model output from the communities for which extensive phylogenetic data were available; this excluded the entire South African component of the study as well as one triplet of communities in Zambia for which health-care facility phylogenetic data were not collected. All the simulations taken forwards for full analysis were from the nine remaining Zambia communities. Transmissions involving individuals in both the “inside” and “outside” patches of the simulation were included (47).

### Phylogenetics

The HPTN 071-02 (PopART) Phylogenetics Study and the sequencing procedure used have been described previously (45,46,50). In summary: samples for HIV genome sequencing were obtained from HIV-positive individuals in the PC (during the earliest visit in which they tested positive for the virus, which could be baseline), and individuals who had been approached at the health-care facilities between 2016 and 2018. These individuals were not on ART at the time they were recruited and were in most attending the clinics for ART initiation, although some self-reported previous ART and poor adherence. Whole-genome deep sequencing was done on the Illumina platform using veSEQ-HIV (60).

The process by which probable transmission pairs were identified has also been previously described (50). Briefly, after genome assembly using SPAdes (61) and *shiver* (38), the collection of viral consensus sequences was clustered using HIV-TRACE (28) with an intentionally generous distance threshold of 0.04 substitutions per site. The deep-sequence data files from all members of every identified cluster were then used as input for separate analyses with *phyloscanner* v1.8.0 (27). The set of genomic windows used in *phyloscanner* were uniformly of 250bp in length, spanning the entire HIV genome, such that the start of each window was 10 bp further along the genome (relative to HXB2) than the previous one. The distance threshold used to identify likely transmission pairs in a single window was of 0.025 substitutions per site on a normalised scale designed to represent average distances on the *gag* and *pol* genes (62). After adjustment for missing windows, overall assignment of pairs as likely to involve direct transmission was declared where more than 50% of windows suggested it. Same-sex pairs (which indicate at least one missing intermediary under the assumption that transmission is heterosexual) were then discarded to arrive at a final set; previous work has estimated that the proportion of true male-to-male transmissions was very low amongst trial participants (50).

The date of HIV infection of each individual was estimated using HIV-phyloTSI (63) and these dates, together with recorded dates of birth, were used to estimate the age of source and recipient partners in each pair at the time of the infection of the recipient. While our previous analysis (50) investigated only transmissions estimated to have occurred during the time-frame of the PopART trial itself, this filter was not applied here.

Direction of transmission was determined using both the *phyloscanner* score based on the tree topology and comparison of infection dates as estimated by HIV-phyloTSI; see (50) for full details.

### Posterior predictive test for phylogenetics results against the IBM

To test whether the phylogenetics results were compatible with being derived from the IBM, we performed a posterior predictive check. This determines the probability that a particular statistical estimate from the phylogenetics (in this case the mean source or recipient age, or mean age gap) is drawn from the posterior distribution of that statistic from the IBM. We accounted for error in the phylogenetics estimate by, for each posterior draw, also drawing an error term from a normal distribution with mean 0 and standard deviation equal to the standard error of the phylogenetics estimate.

## Appendix 1 Comparison of identified pairs from phylogenetics to random pairs

In this appendix, we explore the presence of a genuine signal of HIV transmission in the phylogenetics dataset, by comparing the results obtained from *phyloscanner* to the results of attaching two opposite-sex individuals in the genomic dataset to each other at random as false transmission pairs.

Specifically, we picked a random selection of 156 male and 197 female participants for whom sequence data were available. (156 and 197 are the number of recipients of each sex in the actual dataset.) We additionally selected these such that their estimated infection dates (as determined by phyloTSI) were no earlier than the first such estimate amongst the recipients in the actual data. For each of these 353 individuals we randomly picked an individual of the opposite sex, whose estimated infection date was earlier than their own, as a shuffled “source” for their transmission. The time of transmission was taken to be the estimated time of infection of the “recipient”. We repeated this process 1,000 times and compare the distribution of age- and sex-related variables across these replicates to those in the real dataset (Appendix 1-Figures 1 to 3).

### Recipients

See Appendix 1-Figure 1 for a comparison of the mean and variance of recipients age at the time of transmission, by sex, across the replicates of the shuffled dataset (histograms) to its true value (green line). There is no evidence of a difference with respect to either mean or variance for either recipient sex (all empirical *p-*values >0.178). This provides reassurance that the age distribution of individuals with a sampled source does not differ from that of the complete set of participants providing sequences.

Estimated time since infection showed considerable variation between real and shuffled datasets (Appendix 1-Figure 2), with individuals from shuffled pairs (i.e. those whose sources had not genuinely been sampled) showing greater mean values of the variable (empirical *p*-values <0.001 for both sexes). This is consistent with younger infections being more likely to have a sampled source; elapsed time from infection would be associated with increasing probability that the source would not be eligible for sampling, either due to migration or commencing ART (as successful ART treatment prevents the acquisition of a sequence).

### Sources

In appendix 1-figure 3, mean estimated source ages at the time of transmission are compared. For male sources there is no evidence of a difference in mean age (empirical *p*=0.41 but female sources are younger in the actual dataset (*p*<0.001). The variances of source ages also deviate from the null hypothesis for both sexes (*p*=0.02 for males, *p*<0.001 for females). Effectively, this test compares the demographics of those individuals identified as sources by the analysis to the general HIV positive population, as the shuffled “sources” need not have genuinely transmitted HIV. It is unsurprising that these variables differed. In this case, a younger age for female sources of transmission when compared to a random sample of HIV-positive women is consistent with onward transmission peaking in younger age groups. A smaller variance in the ages of genuine sources is also consistent with their being a peak age of risk for transmitting the virus (see figure 1).

As with recipients, “sources” in the shuffled dataset tend to have older infections (Appendix 1-Figure 4). In this case this is likely due to a combination of the same rationale outlined for recipients above, and the fact that genuinely identified sources are younger than a random selection of individuals (empirical *p*<0.001 for both sexes). They would be expected, on average, to have more recent infections because the range of possible times since infection is constrained by a more recent sexual debut.

### Age gap

Appendix 1-Figure 5 displays the histograms for the age gap between male and female partners. There is strong evidence for a difference in means for female-to-male transmission (empirical *p*<0.001) but not for male-to-female (*p*=0.307). Meanwhile there are plainly large differences in the variances (*p*<0.001), meaning that genuine transmission pairs have much more closely correlated ages than random pairs.

## Discussion

The sampling frame for this study is unusual. We have taken a sample of HIV-positive individuals in the trial communities, and within that sample identified individuals who we are confident acted as sources of transmission for others in the sample. Only 4.96% of sampled individuals have a source identified in this way. As every HIV-positive individual is by definition a recipient of transmission, this 4.96% can be seen as a subsample of the sampled recipient population. If the subsample were unrepresentative, meaning that those recipients for whom we found a source were different to those for whom we did not, it would in turn suggest bias in the characteristics of the sources that we identified for these recipients. That there was no statistical difference between the true subsample of recipients with sampled sources and random collections of recipients of the same size should give us confidence that this bias was not present.

The *phyloscanner* procedure would obviously have failed if it was doing no more than sampling random opposite-sex pairs of individuals in the dataset and designating one of each as the source of the infection in the other. Were this true, then the demographics of the population identified as sources would be no different from those of the entire sample, and the distribution of age gaps in identified pairs would be no different from that of random pairs of opposite-sex individuals in the dataset. We found strong statistical evidence that neither of these are the case.

## Appendix 2 Sensitivity analysis for *phyloscanner* settings

In this section we re-investigate the age and sex dynamics of the set of identified transmission pairs from sets of pairs obtained when some of the key *phyloscanner* settings are varied. For further information regarding these settings, see the *phyloscanner* paper (27) and manual (available at https://github.com/BDI-pathogens/phyloscanner)

### Read downsampling

The nature of deep sequencing is such that genome coverage will vary both between samples and in different regions of the genome for the same sample. It may well be the case that, for a given *phyloscanner* window and a putative transmission pair, the number of reads covering the window can differ between the two members of the pair. The exact effects of this on inference have yet to be exhaustively explored, but one *phyloscanner* option allows the user to specify a maximum number of reads to be analysed per window, with the exact choice of reads being randomly selected. (Note that, as many reads in the same window will be identical due to the amplification process during sequencing, regardless of underlying variation within the sample, this process is not the same as limiting the number of distinct genomic *variants*.) A further choice must be made regarding how to proceed if the total number of reads for one host in one window fails to reach that maximum number.

The two options are, firstly, to include all the reads that do exist (“keep”) or, secondly, to remove that host from consideration entirely in that window (“blacklist”). (As no inference is conducted from windows in which reads are not present from both hosts, the practical effect of the “blacklist” option is that those windows will be excluded from the analysis.) In the main analysis of this paper we did not use read downsampling; here we describe results from applying maximum read counts of 25 and 50, with both the “keep” and “blacklist” strategies.

As described in the main text, there are 355 pairs found when no read downsampling was applied. Applying the tool always reduces this number, to:

● 296 with a maximum of 50 reads per window and the “keep” strategy
● 274 with a maximum of 50 reads per window and the “blacklist” strategy
● 285 with a maximum of 25 reads per window and the “keep” strategy
● 284 with a maximum of 25 reads per window and the “blacklist” strategy

There was no evidence of a difference in the proportion of pairs with a male source (chi-squared test *p*=0.699) between the five groups. In this work, we have three key variables of interest: source age at estimated time of transmission, recipient age at estimated time of transmission, and age gap. Each of these was examined separately according to the sex of the inferred source individual, making a total of six variables in practice. We saw no evidence of a difference in any of these variables regardless of the downsampling procedure. (Kruskall-Wallis test *p*>0.7 in all cases). Their distribution can be seen in appendix 2-figure 1.

### Window width

A key *phyloscanner* setting is the width of the sliding window used to subdivide the viral genome. Narrower windows allow the genome to be tiled with more windows and increase the number of reads per window because *phyloscanner* requires reads to fully span a window to be included in it. However, narrow windows have fewer sites at which mutations can be observed, reducing phylogenetic resolution. In the main text the window width was set to 250 base pairs, with each window starting 10bp further along than the previous. We also reduced this to 175bp, with the same 10bp incremental shift, and obtained the set of probable transmission pairs from this variation. The 355 pairs from the main analysis were reduced to 291 here. There was no evidence that these sets differed in respect to the proportion of pairs which were male-to-female (chi-squared test *p*=0.903) or of our six key quantities (Mann-Whitney *U*-test *p*>0.63 in all cases). See appendix 2-figure 2 for the distribution of these values.

### Window thresholds for pair identification

As phylogenetic relationships between the reads from each of two individuals will vary across the genome, we apply thresholds on the proportion of windows in which a) the phylogeny suggests that the pair are a transmission pair (“linkage”), and b) the phylogeny suggests a direction of transmission. (These proportions are adjusted for non-informative windows as outlined by (62)). In the main analysis, individuals are linked as a pair at a proportion threshold of 0.5. For pairs that are above this threshold, a direction of transmission is called at a proportion threshold of 0.33 (of the total number of windows). We also tested more stringent thresholds, firstly 0.66 for linkage and 0.5 for direction, and then 0.75 for linkage and 0.66 for direction. The 355 pairs from the main analysis were reduced to 260 in the former case and 201 in the latter. There is no evidence of a difference in the proportion of pairs identified as having a male source (chi-squared test *p*=0.927) or in the distribution of our six quantities of interest (Kruskal-Wallis test, all *p*>0.3). See appendix 2-figure 3 for the distribution of these estimates.

### Patristic distance threshold for per-window pair identification

The final setting that we varied was the patristic distance threshold at which, in a single genomic window, a pair of individuals are inferred as likely to be involved in direct transmission. As phylogenetic branch lengths vary across the genome, this distance is normalised to standardise to expected lengths on the *gag* and *pol* genes. In the main analysis the value used is 0.02 normalised substitutions per site. Here we use values from 0.01 to 0.04 in increments of 0.005.

Increasing this threshold should increase sensitivity, but decrease specificity, of the identification of transmission pairs. For heterosexual HIV transmission, instances where there is only a single unsampled intermediate host can be omitted by simply excluding same-sex pairs (e.g. male-male pairs must be missing at least one unsampled intermediate female). Then false positives must be missing at least two unsampled intermediaries (for example identifying male A → female B → male C → female D as male A → female D). Reducing the patristic distance threshold will reduce the probability that such false positives are included in the dataset. See appendix 2-table 1 for the number of pairs identified using each threshold.

There is no evidence of an association of the threshold with the proportion of transmission pairs identified with male sources, whether the threshold is treated as a discrete variable (chi-squared test *p*=0.962), or as continuous (logistic regression *p*=0.376). Similarly, none of our six key estimates show evidence for an association with the threshold. Again, this is true whether the threshold is treated as a discrete variable (Kruskal-Wallis *p*>0.8 in all cases) or fit as a continuous predictor of the variable in a linear regression model, with source sex as an interaction term (no coefficients other than source sex having *p*<0.3). See appendix 2-figure 4 for the actual distribution of their values.

That the key outputs of our analysis are robust to decreasing the distance threshold from 0.02 to 0.01 should offer reassurance towards concerns regarding any biasing effect on the results of this paper caused by the inclusion of false positive transmission pairs.

## Data Availability

For the genomic data, consensus sequences with PANGEA-specific identifiers, sex of participant, and year and country of sampling are available at https://github.com/PANGEA-HIV/PANGEA-Sequences. The analysis code will also be made available on Zenodo upon publication of the version of record. Additional data fields and next-generation sequencing data can be requested via the data sharing procedures of the PANGEA consortium (see www.pangea-hiv.org). The requests are assessed by the PANGEA Steering Committee and data will usually be made available for non-commercial purposes unless there is a risk of participants and communities being harmed or stigmatised, or participants' privacy being compromised. If multiple applicants are aiming to pursue the same analyses, they will be encouraged to coordinate efforts. IBM code is fully open source and the code necessary to run it is available at https://github.com/BDI-pathogens/hiv-phylo-age-sex.

http://github.com/BDI-pathogens/hiv-phylo-age-sex

---

**Appendix 2-Table 1.** Number of phylogenetic transmission pairs identified using different patristic distance thresholds.

| Distance threshold (substitutions per site, normalised to represent average distances on the *gag* and *pol* genes) | Number of pairs identified |
| --- | --- |
| 0.01 | 257 |
| 0.015 | 315 |
| 0.02 | 355 |
| 0.025 | 386 |
| 0.03 | 417 |
| 0.035 | 441 |
| 0.04 | 459 |

**Appendix 1-Figure 1:**
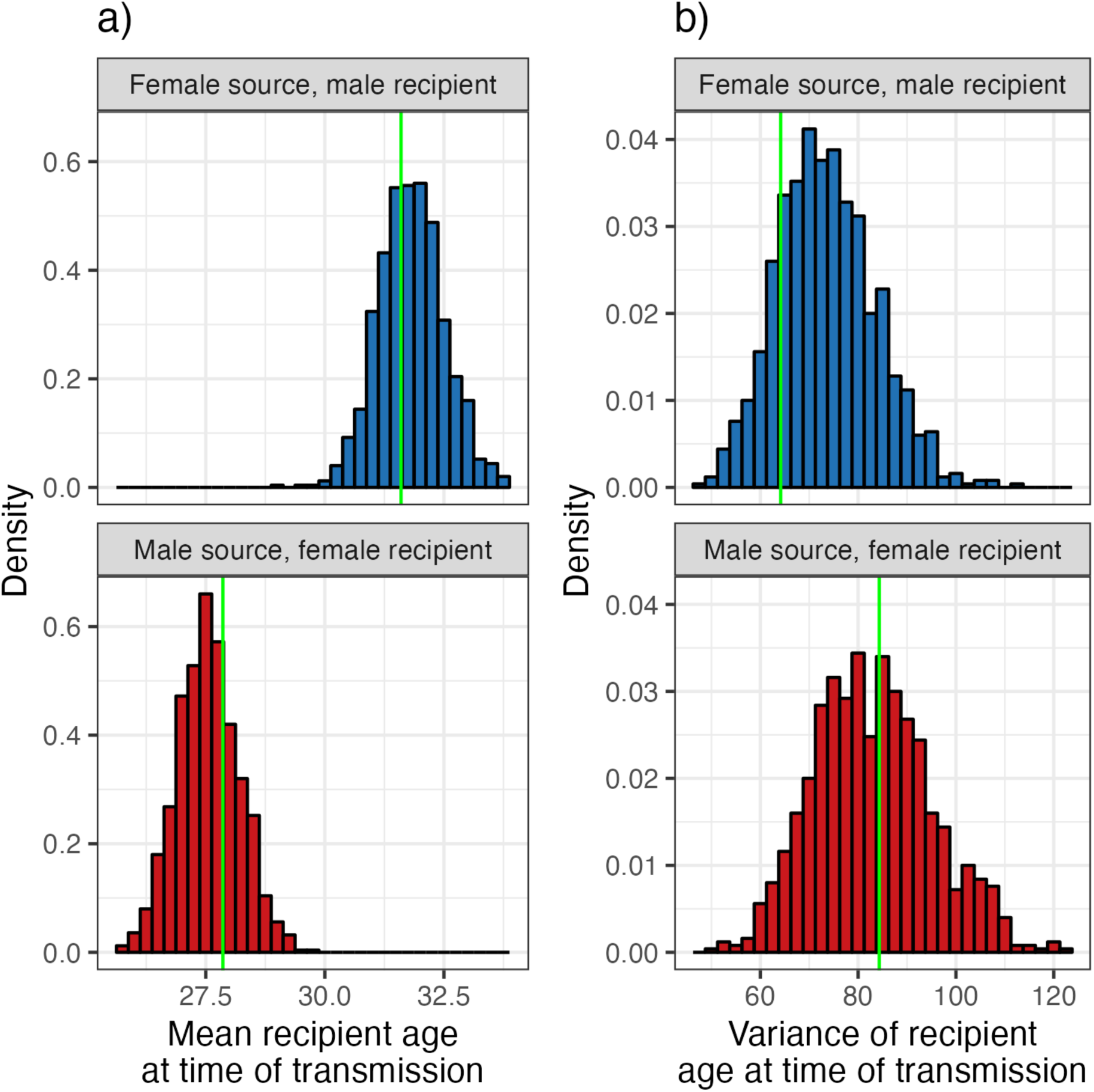
Empirical distributions of mean (a) and variance (b) of ages of recipients at time of transmission from pairs where “recipients” had randomly chosen “sources” (histograms) compared to the true value from the phylogenetics data (green line).

**Appendix 1-Figure 2:**
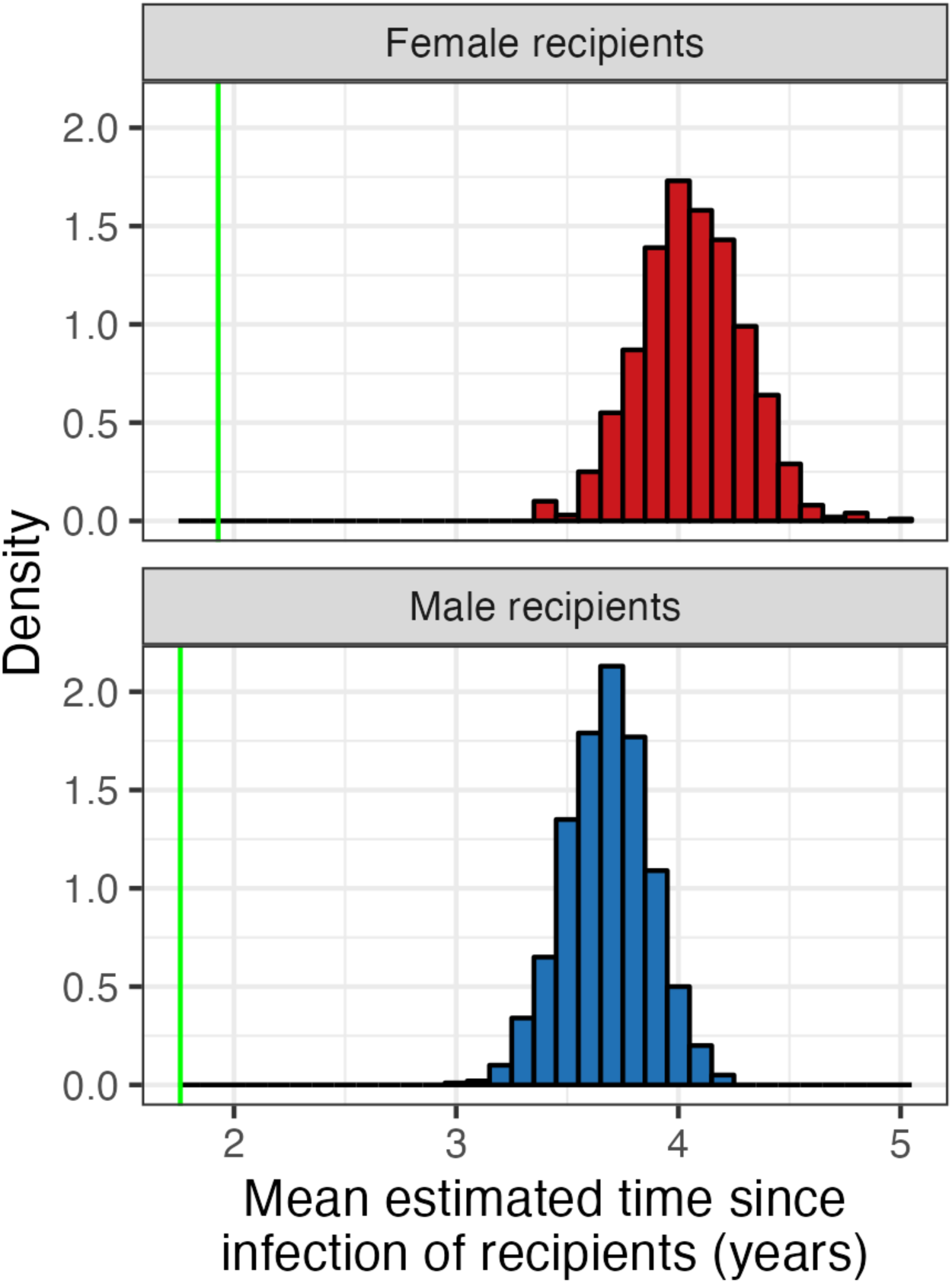
Empirical distributions of mean estimated time from infection to sampling of recipients from randomised transmission pairs (histograms) compared to the true value from the phylogenetics data (green line).

**Appendix 1-Figure 3:**
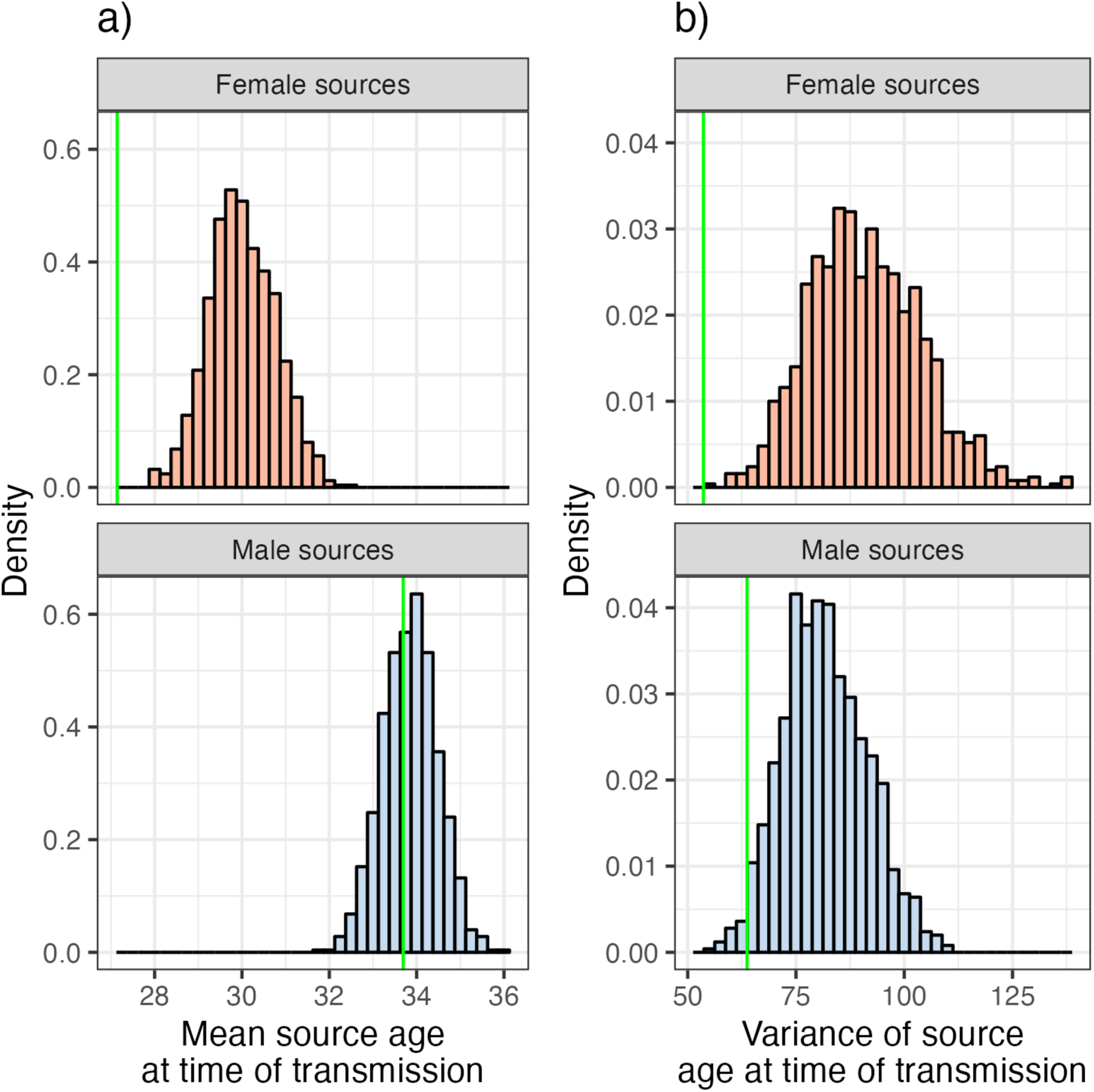
Empirical distributions of mean (a) and variance (b) of ages of sources at time of transmission from randomised transmission pairs (histograms) compared to the true value from the phylogenetics data (green line).

**Appendix 1-Figure 4:**
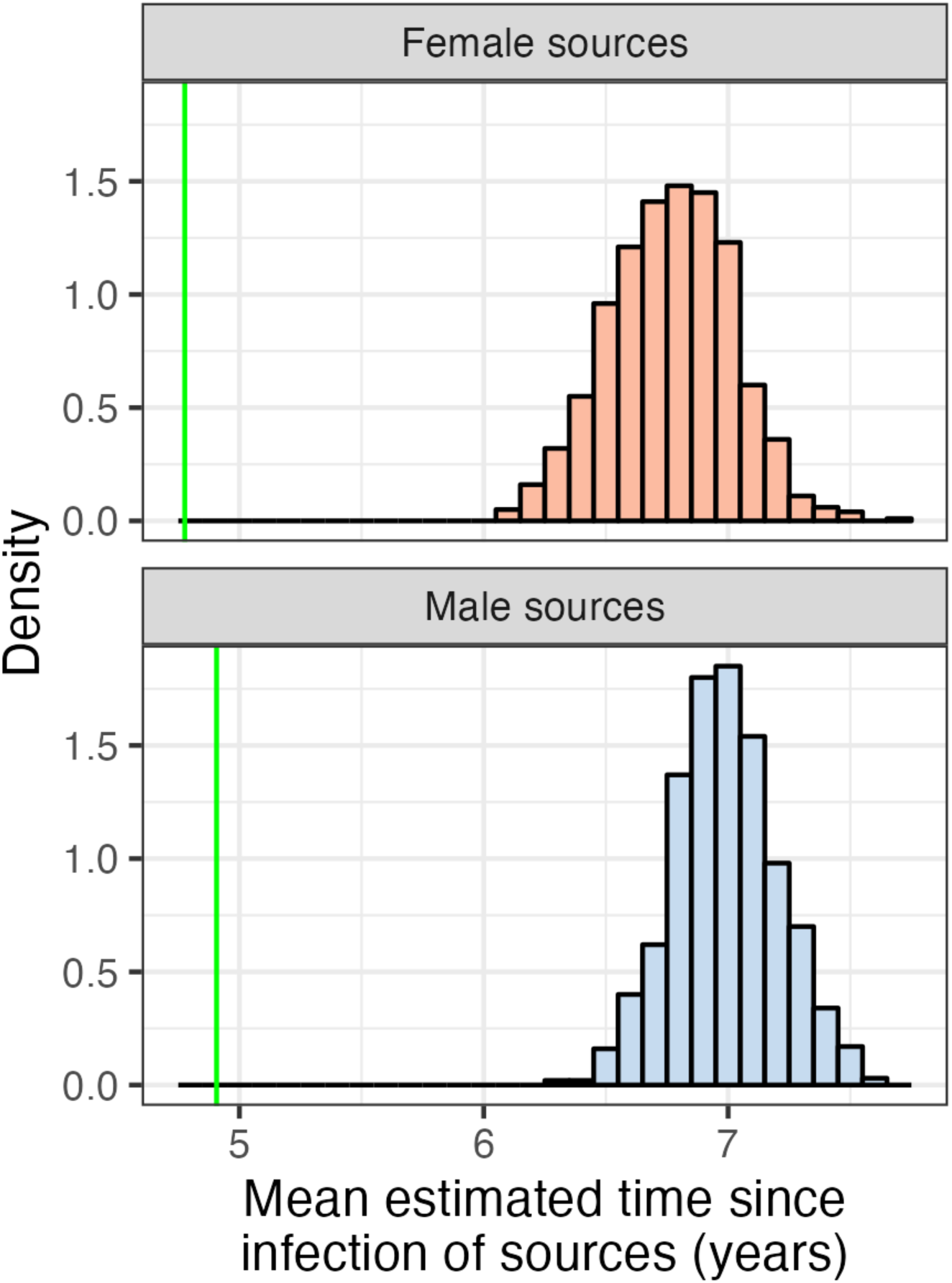
Empirical distributions of mean estimated time from infection to sampling of sources from randomised transmission pairs (histograms) compared to the true value from the phylogenetics data (green line).

**Appendix 1-Figure 5:**
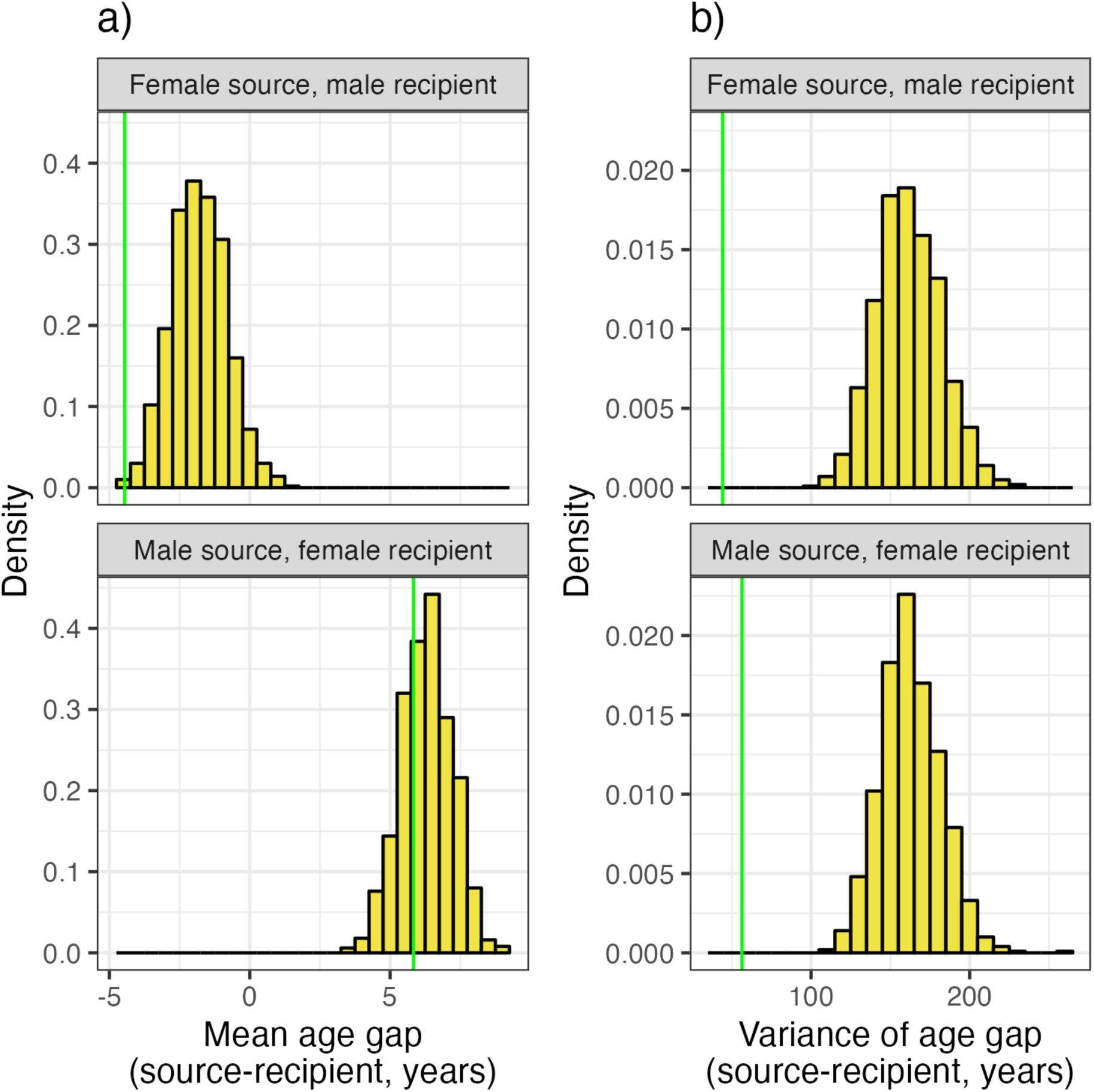
Empirical distributions of mean (a) and variance (b) in transmission age gap from randomised transmission pairs (histograms) compared to the true value from the phylogenetics data (green line).

**Appendix 2-Figure 1:**
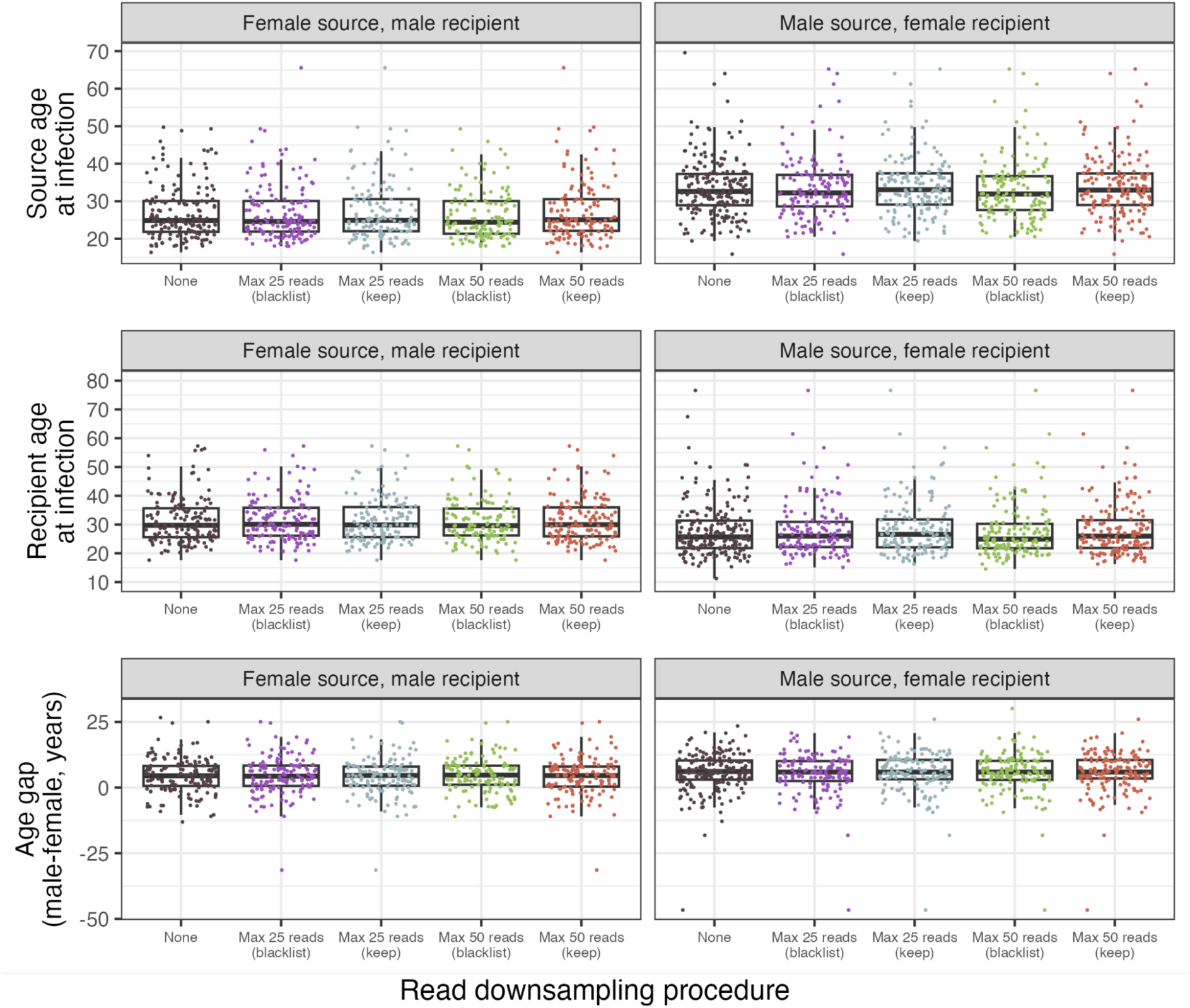
Distributions of the estimated ages of sources at the time of transmission (top), estimated ages of recipients at the time of transmission (middle) and age gap amongst probable transmission pairs identified using *phyloscanner* when the read downsampling procedure is varied. Downsampling to a maximum of *n* reads means that a random sampling of *n* reads is taken for each individual in each genomic window. For windows in which *n* reads do not exist, the two options are “blacklist” - to exclude that host from that window entirely - and “keep” - to use all the reads that are available. The boxplots display the median, 25% and 75% quantiles, and range.

**Appendix 2-Figure 2:**
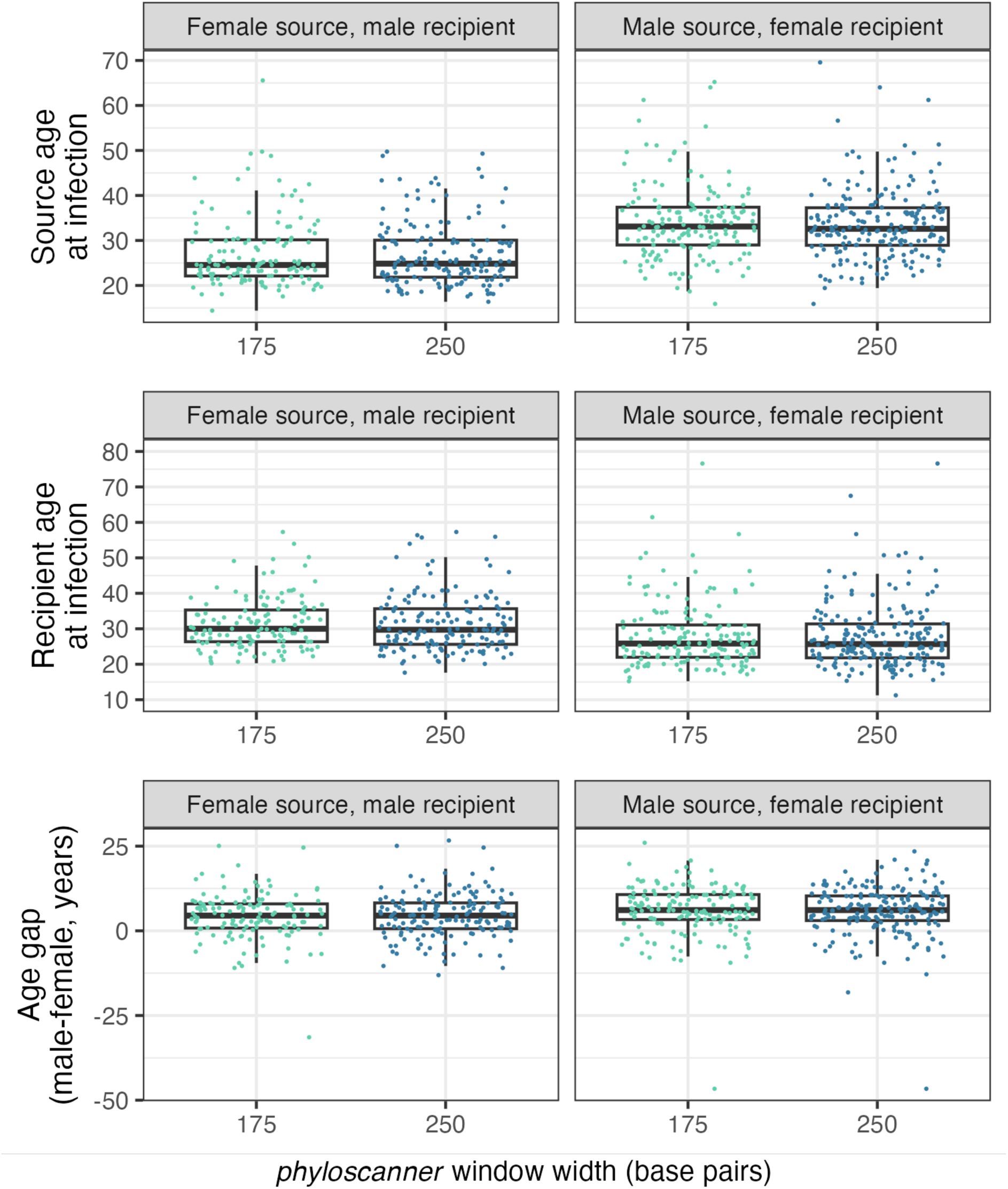
Distributions of the estimated ages of sources at the time of transmission (top), estimated ages of recipients at the time of transmission (middle) and age gap amongst probable transmission pairs identified using *phyloscanner* with different genomic window widths. The boxplots display the median, 75% quantiles, and range.

**Appendix 2-figure 3:**
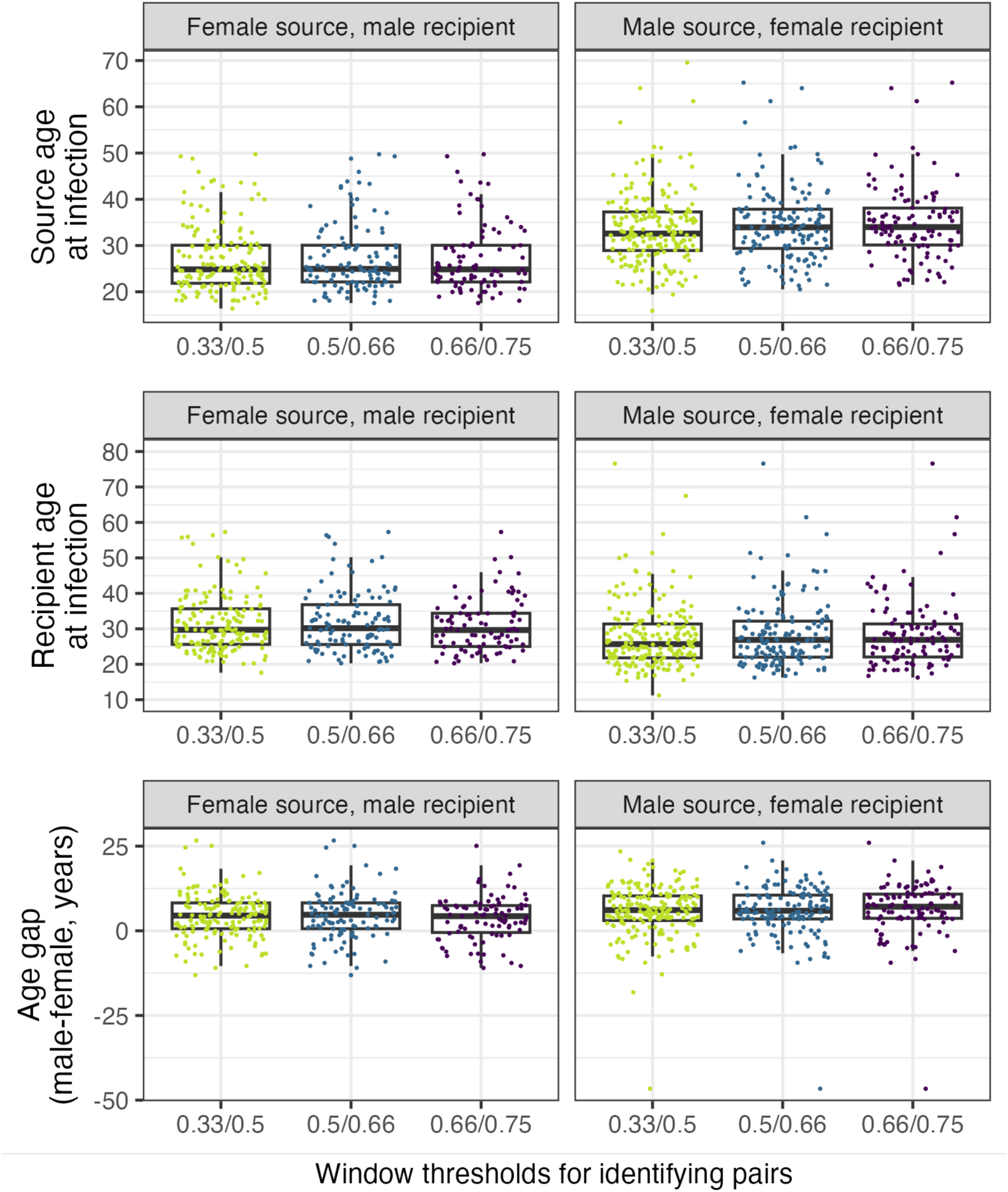
Distributions of the estimated ages of sources at the time of transmission (top), estimated ages of recipients at the time of transmission (middle) and age gap amongst probable transmission pairs identified using *phyloscanner*. Different window-related thresholds for identifying linkage and direction are indicated on the x axis in the format X/Y, where Y is the minimum fraction of genomic windows supporting two samples being from a transmission pair, and X is the minimum fraction of genomic windows supporting transmission having an identifiable direction (conditional on being a transmission pair). The boxplots display the median, 75% quantiles, and range.

**Appendix 2-Figure 4:**
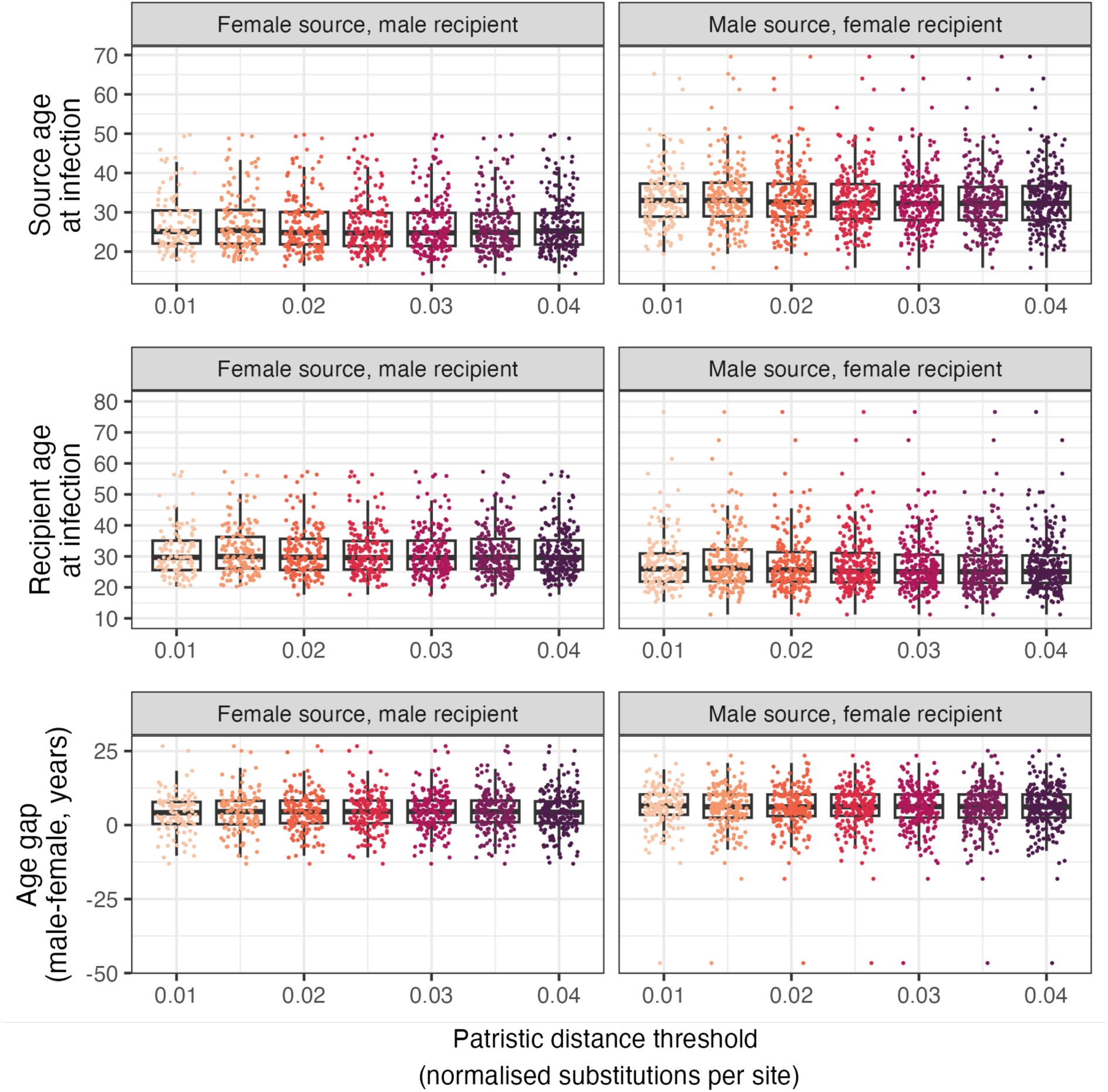
Distributions of the estimated ages of sources at the time of transmission (top), estimated ages of recipients at the time of transmission (middle) and age gap amongst probable transmission pairs identified using *phyloscanner* with patristic distance thresholds used to identify transmission pairs in an individual window. The boxplots display the median, 75% quantiles, and range.

## Notes

### Competing Interest Statement

The authors have declared no competing interest.

### Funding Statement

This work was supported by the National Institute of Allergy and Infectious Diseases, the US President's Emergency Plan for AIDS Relief, the International Initiative for Impact Evaluation, the Bill and Melinda Gates Foundation, the National Institute on Drug Abuse, and the National Institute of Mental Health. The content herein is solely the responsibility of the authors and does not necessarily represent the official views of the funding agencies. TG is supported by an Investigator Grant (GNT2025445) from the National Health and Medical Research Council, Australia (NHMRC). MP and AC are supported by the HIV Prevention Trial Network (HPTN) Modelling Centre, funded by the US National Institutes of Health (grant number UM1-AI068617)

### Author Declarations

The National Institutes of Health Institutional Review Board gave ethical approval of this work The Biomedical Research Ethics Committee of the University of Zambia gave ethical approval of this work The Observational/Interventions Research Ethics Committee of the London School of Hygiene and Tropical Medicine gave ethical approval of this work

### Summary of Updates

The authors list has been corrected, and minor changes to the text made after author comments in preparation for eLife resubmission.

